# Multidimensional Data Integration Identifies Tumor Necrosis Factor Activation in Nephrotic Syndrome: A Model for Precision Nephrology

**DOI:** 10.1101/2021.09.09.21262925

**Authors:** Laura H. Mariani, Sean Eddy, Fadhl M. AlAkwaa, Phillip J. McCown, Jennifer L. Harder, Sebastian Martini, Adebowale D. Ademola, Vincent Boima, Heather N. Reich, Felix Eichinger, Jamal El Saghir, Bradley Godfrey, Wenjun Ju, Viji Nair, Emily Tanner, Virginia Vega-Warner, Noel L. Wys, Sharon G. Adler, Gerald B. Appel, Ambarish Athavale, Meredith A. Atkinson, Serena M. Bagnasco, Laura Barisoni, Elizabeth Brown, Daniel C. Cattran, Katherine M. Dell, Vimal K. Derebail, Fernando C. Fervenza, Alessia Fornoni, Crystal A. Gadegbeku, Keisha L. Gibson, Larry A. Greenbaum, Sangeeta R. Hingorani, Michelle A. Hladunewich, Jeffrey B. Hodgin, Jonathan J. Hogan, Marie Hogan, Lawrence B. Holzman, J. Ashley Jefferson, Frederick J. Kaskel, Jeffrey B. Kopp, Richard A. Lafayette, Kevin V. Lemley, John C. Lieske, Jen-Jar Lin, Rajarasee Menon, Kevin E. Meyers, Patrick H. Nachman, Cynthia C. Nast, Alicia M. Neu, Michelle M. O’Shaughnessy, Edgar A. Otto, Kimberly J. Reidy, Kamalanathan K. Sambandam, John R. Sedor, Christine B. Sethna, Pamela Singer, Tarak Srivastava, Cheryl L. Tran, Katherine R. Tuttle, Suzanne Vento, Chia-shi Wang, Akinlolu O. Ojo, Dwomoa Adu, Debbie S. Gipson, Howard Trachtman, Matthias Kretzler

## Abstract

**Background:** Classification of nephrotic syndrome relies on clinical presentation and descriptive patterns of injury on kidney biopsies. This approach does not reflect underlying disease biology, limiting the ability to predict progression or treatment response.

**Methods:** Systems biology approaches were used to categorize patients with minimal change disease (MCD) and focal segmental glomerulosclerosis (FSGS) based on kidney biopsy tissue transcriptomics across three cohorts and assessed association with clinical outcomes. Patient-level tissue pathway activation scores were generated using differential gene expression. Then, functional enrichment and non-invasive urine biomarker candidates were identified. Biomarkers were validated in kidney organoid models and single nucleus RNA-seq (snRNAseq) from kidney biopsies.

**Results:** Transcriptome-based categorization identified three subgroups of patients with shared molecular signatures across independent North American, European and African cohorts. One subgroup demonstrated worse longterm outcomes (HR 5.2, p = 0.001) which persisted after adjusting for diagnosis and clinical measures (HR 3.8, p = 0.035) at time of biopsy. This subgroup’s molecular profile was largely (48%) driven by tissue necrosis factor (TNF) activation and could be predicted based on levels of TNF pathway urinary biomarkers TIMP-1 and MCP-1 and clinical features (correlation 0.63, p <0.001 for predicted vs observed score). Kidney organoids confirmed TNF-dependent increase in transcript and protein levels of these markers in kidney cells, as did snRNAseq from NEPTUNE biopsy samples.

**Conclusions:** Molecular profiling identified a patient subgroup within nephrotic syndrome with poor outcome and kidney TNF pathway activation. Clinical trials using non-invasive biomarkers of pathway activation to target therapies are currently being evaluated.

**Significance Statement:** Mechanistic, targeted therapies are urgently needed for patients with nephrotic syndrome. The inability to target an individual’s specific disease mechanism using currently used diagnostic parameters leads to potential treatment failure and toxicity risk. Patients with focal segmental glomerulosclerosis (FSGS) and minimal change disease (MCD) were grouped by kidney tissue transcriptional profiles and a subgroup associated with poor outcomes defined. The segregation of the poor outcome group was driven by tumor necrosis factor (TNF) pathway activation and could be identified by urine biomarkers, MCP1 and TIMP1. Based on these findings, clinical trials utilizing non-invasive biomarkers of pathway activation to target therapies, improve response rates and facilitate personalized treatment in nephrotic syndrome have been initiated.

## Introduction

Nephrotic syndrome refers to kidney diseases marked by proteinuria, hypoalbuminemia, hyperlipidemia and edema. Glomerular diseases associated with this constellation of clinical features include minimal change disease (MCD) and focal segmental glomerulosclerosis (FSGS). These disease entities are currently classified based on distinct histopathological features observed in kidney biopsies. However, for a given histopathologic diagnosis, the clinical presentation may vary considerably (e.g. rapidity of onset and degree of edema); conversely, distinct histopathologic diagnoses might share clinical features, reflecting heterogeneity and poor understanding of underlying biological processes ^1–3^. Clinical decisions rely on these histopathologic categories, combined with routine clinical parameters (e.g. serum creatinine and urine protein) and response to initial empiric therapy (e.g. steroid sensitive vs. resistant disease). However, due to the diagnostic imprecision and biological limitations of descriptive disease classification, molecularly targeted treatments for MCD and FSGS are not available. Further, inclusion of heterogeneous patient populations challenges the interpretation of results from observational studies and clinical trials of therapeutic agents ^4^.

Precision medicine for glomerulopathies can be enabled through recent advances in biomedical research that allow capture of data domains across the genotype-phenotype continuum from patients under routine clinical care ^5, 6^. This approach integrates data across multiple domains, and pairs it with in-depth phenotyping to establish a novel disease classification reflecting distinct molecular states of the patients. The goal of this approach is to improve prediction of progression risk and discover safer and more effective targeted treatments. In particular, targeted therapy trials can be enriched for participants affected by specific molecular pathways^7^.

Nephrotic syndrome is uniquely positioned to implement this approach. Clinically procured kidney biopsy tissue allows for identification of molecular signatures that can then be linked to detailed histopathology, non-invasive biomarkers and evaluated with clinical outcomes. In this study, we implement these approaches (Figure 1) in the prospective North-American Nephrotic Syndrome Study Network (NEPTUNE) as the discovery cohort and replicate our findings in the European Renal cDNA Bank (ERCB) the Human Heredity and Health in Africa Kidney Disease Research Network cohort (H3Africa).

**Figure 1:**
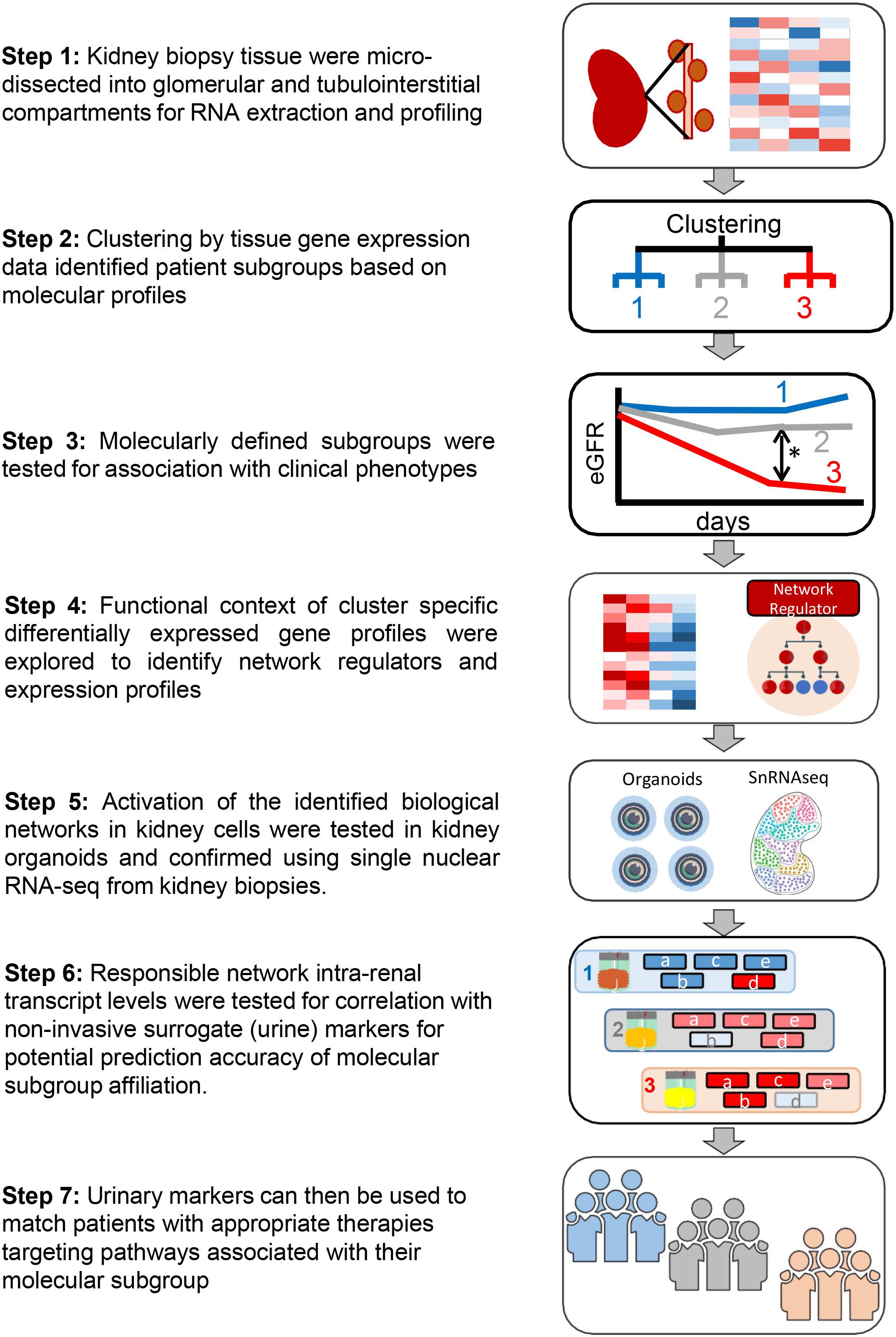
Analysis strategy. Flowchart of tubulointerstitial compartment gene expression to identify molecular subgroups and associated non-invasive urinary markers.

## Methods

### Study Participants

The study involved 220 participants with biopsy-proven MCD or FSGS enrolled in the prospective NEPTUNE study^8^, 35 participants with biopsy-proven MCD or FSGS enrolled in the H3Africa study^9^, and 30 participants with biopsy-proven MCD or FSGS from the ERCB^10, 11^. Participants with compartment-enriched genome-wide kidney mRNA expression profiles of their kidney biopsies were included.

NEPTUNE (NCT01209000) is a multi-center (21 sites), prospective study of children and adults with proteinuria (>500mg/day in phase I and 1.5g/day in phase 2), recruited at the time of first clinically indicated kidney biopsy. It was launched in August 2010. The objectives, study design and procedures have been described in detail in previous publications^8, 12^. ERCB is a European multicenter study that collects biopsy tissue for gene expression profiling along with cross-sectional clinical information, e.g., demographics, eGFR (estimated glomerular filtration rate), at the time of a clinically indicated kidney biopsy in adults across 28 sites^10, 11^. The subset of participants with MCD or FSGS were included in the validation cohort for the gene expression analyses.

The H3 Africa cohort study^9^ is a multi-center, prospective study of patients aged 15 years and above, recruited from 13 participating clinical centers in Nigeria and Ghana. Participants with eGFR ≥15 and proteinuria (albuminuria >500mg/day) were eligible for a kidney biopsy and composed the glomerulonephritis arm of the study. The study design has been described previously^9^.

For all cohorts, consent was obtained from individual patients or parents/guardians at enrollment, and the studies were approved by Institutional Review Boards or local ethics committees of participating institutions. NEPTUNE was approved (HUM00158219) by University of Michigan, Medical School Institutional Review Board. The ERCB ethics committee approval from the Ethikkommission bei der LMU Munchen, Ludwig-Maximilian-Universitat Munchen, Pettenkoferstr. 8a, 80336 Munchen, Ethical approval under 250-16. The H3 Africa study, STUDY00144768, was approved by University of Kansas Medical Center Institutional Review Board and ethics committees at each clinical site: Lagos University Teaching Hospital Health Research Ethics Committee (ADM/DCST/HREC/APP/1550); Aminu Kano Teaching Hospital (AKTH / MAC / SUB / 12A / P-3 / VI / 2032); Lagos State University Teaching Hospital (REF NO: LREC. 06/10/933); University Of Abuja Teaching Hospital (REF NO: FCT/UATH/HREC/PR/554); Ghana Health Service Ethics Review Committee (GHSERC: 014/07/19); Delta State University Teaching Hospital Health Research Ethics Committee (DELSUTH/HREC/2016/050/0198); Usmanu Danfodiyo University Teaching Hospital Health Research Ethics Committee (UDUTH / HREC / 2017 / 594); Nnamdi Azikiwe University Teaching Hospital (NAUTH/CS/66/VOL.12/ 058/2019/039); Kwame Nkrumah University of Science and Technology College Of Health Sciences Committee on Human Research, Publication and Ethics (Ref: CHRPE/ AP /335/20); Obafemi Awolowo University Teaching Hospital (ERC/2017 /06/16); University of Ibadan, College of Medicine (UI/EC/16/0399); University Of Nigeria Teaching Hospital (UNTH/CSA/329/VOL.5/010); University Of Ilorin Teaching Hospital (UITH ERC Protocol number: ERC PIN/2017/01/0511, UITH ERC Approval number: ERC PAN/2020/12/0107).

### Clinical Data

NEPTUNE participants were followed prospectively, every 4 months for the first year, and then biannually thereafter for up to 5 years. At each study visit, medical history, medication use, and standard local laboratory test results were recorded, while blood and urine specimens were collected for central measurement of serum creatinine and urine protein/creatinine ratio (UPCR). eGFR (mL/min/1.73m^2^) was calculated using the CKD-Epi formula for participants ≥18 years old and the modified CKiD-Schwartz formula for participants <18 years old, with an average of the two results taken for adolescents ^13–15^. ESKD was defined as initiation of dialysis, receipt of kidney transplant or eGFR <15 mL/min/1.73m^2^ measured at two sequential clinical visits; and the composite endpoint of kidney functional loss by a combination of ESKD or 40% reduction in eGFR^16^. Complete remission was defined as UPCR <0.3 mg/mg on a single void specimen or 24-hour urine collection.

In ERCB, clinical information, including demographics and local lab results, were recorded at time of biopsy. Similarly, in the H3Africa study patient demographics, serum creatinine, and urine albumin:creatinine ratio were obtained at time of biopsy.

### Interstitial Fibrosis and Tubular Atrophy

In NEPTUNE, the degree of interstitial fibrosis (IF), and tubular atrophy (TA) were visually assessed by pathologists using whole slide imaging of all available biopsy slides using trichrome, PAS, or silver-stained sections. Estimated percent of cortex involved by IF or TA was highly concordant and reproducible across pathologists^17^.

### Transcriptome profiling

In NEPTUNE, the research core obtained at the time of a clinically-indicated biopsy was placed in RNA preservative (RNAlater). Genome wide transcriptome analysis was performed on manually micro-dissected kidney biopsy tissue that separated the tubulointerstitial compartment from the glomerular compartment. For RNA-sequencing (RNA-seq) profiles, mRNA samples were prepared using the Illumina TruSeq mRNA Sample Prep v2 kit. Multiplex amplification was used to prepare cDNA with a paired-end read length of 100 bases using an Illumina HiSeq2000. RNAseq was performed by the University of Michigan Advanced Genomics Core (https://brcf.medicine.umich.edu/cores/advanced-genomics/). Quality of the sequencing data was assessed using the FastQC tool (http://www.bioinformatics.babraham.ac.uk/projects/fastqc/). Read counts were extracted from the fastq files using HTSeq (version 0.11). RNA-seq profiles from different batches were voom-transformed and batch corrected using ComBat^18^.

For ERCB biopsy samples, total RNA was isolated, reverse transcribed, linearly amplified and hybridized on the Affymetrix microarray platforms ^17, 19–22^. Microarrays were preprocessed and normalized with RMA^23^ following the workflow described by Lockstone et al.^24^ Human microarrays were annotated using custom chip definition files from the University of Michigan (Brainarray), custom chip definition file version 19 ^25, 26^. Normalized gene expression data was batch corrected using ComBat ^27^. Transcriptional profiles of biopsies from patients with MCD and FSGS in the ERCB have been deposited to GEO and are part of accession numbers GSE104954 (tubulointerstitium) and GSE104948 (glomeruli).

For H3 Africa biopsy samples, a 5 mm cortical segment of a kidney biopsy core (beyond that necessary for clinical diagnosis) was placed into an RNA preservative (RNAlater). Manual microdissection was used to separate the tissue into glomeruli and tubulo-interstitial compartments and were processed for RNA profiles as in the NEPTUNE study. NEPTUNE, ERCB and H3 Africa RNAseq gene expression data from microdissected biopsies are available for online interrogation at Nephroseq.org and have been deposited into GEO.

### Cluster analysis, differential expression and functional enrichment analysis

Computational analyses were primarily performed in the R statistical computing environment (R Core Team (2013). R: A language and environment for statistical computing. R Foundation for Statistical Computing, Vienna, Austria (http://www.R-project.org/). Optimal clustering was determined using delta-K and the Consensus Cluster Plus Package^28^. Differential expression analysis was performed using the limma package^29^. Differentially expressed genes (absolute fold change > 1.5 and q-value<0.05) between clusters of interest were analyzed for enrichment of canonical pathways and functional groups using the Ingenuity Pathway Analysis Software Suite (IPA).

Cell-type selective expression clusters were previously published^30^ from single-cell RNA-seq profiles using adult reference kidney tissue. Summary data are deposited in the Adult normal kidney dataset in http://nephrocell.miktmc.org/ and gene expression matrices are found in GEO under the accession number, GSE140989.

### Tumore Necrosis Factor (TNF) Activation Score

A TNF causal network was generated from NetPro expert curated gene effects and interactions in the Genomatix Genome Analyzer database (Precigen Bioinformatics Germany GmbH). From the database, 272 gene symbols representing genes or proteins with increased expression from TNF exposure were used to generate a TNF activation score. In each cohort transcriptomic dataset, individual gene expression values were Z-transformed and the average Z-score of all 272 TNF target genes for each participant was used as that individual’s composite TNF activation score.

### Urine Biomarker Profiling

Biomarkers were identified from a multiplex protein immunoassay data for a panel of 54 urinary cytokines, matrix metalloproteinases and tissue inhibitor of metalloproteinases using the multiplex Luminex platform (Eve Technologies, Alberta, Canada). All urine analyte levels were measured in duplicate and normalized to urine creatinine concentration. Extensive quality control thresholds were required for inclusion in the analysis. Briefly, based on manual review of all analyte distributions, data with technical errors (e.g., low bead count, high inter-well variability) were discarded. Data were flagged if they were extrapolated (outside of the standard curve) or out of range (above or below the 4-5 dilution logistic standard curve). A coefficient of variation was calculated for each analyte aggregated by sample. Analytes were excluded if >50% of measures were extrapolated or out of range or there was a high coefficient of variation. To be evaluated as a potential non-invasive marker of TNF activation: 1) the urine protein had to be a product of a gene causally downstream of TNF; 2) the corresponding intra-renal tissue gene expression (mRNA levels) and 3) the TNF activation score had to correlate with the observed protein levels.

Putative markers identified from the multiplex Luminex platform were then assayed in duplicate from baseline urine specimens using Quantikine ELISA kit Human chemokine (C-C motif) ligand 2 (CCL2) / monocyte chemoattractant protein-1 (MCP-1) (DCP00) and tissue inhibitor of metalloproteinases 1(TIMP-1, DTM100, R&D Systems, Minneapolis, MN, USA). Absorbance was measured with a VersaMax ELISA plate reader, and results were calculated with SoftMax Pro (Molecular Devices). Biomarkers were normalized to urine creatinine concentration and Log2 transformed for analysis.

### Statistical Analysis of the Association with Clinical data and Urine Biomarkers

Descriptive statistics, including mean and standard deviation (SD) for normally distributed variables, median and interquartile range (IQR) for skewed variables and proportions for categorical variables were used to characterize baseline (time of biopsy) participant characteristics by molecular cluster. Kaplan-Meier curves by molecular cluster were generated and overall differences in survival curves tested by the log rank test. Univariate Cox proportional hazard models were fit separately for time to complete remission and time to the composite of end stage kidney disease (ESKD) and loss of 40% decline in eGFR to assess association of molecular cluster and TNF score with clinical outcomes. Patients reaching the endpoint between screening and biopsy visit were excluded from the survival analysis, since their outcome was already known at time of biopsy. Models were adjusted for diagnosis (MCD, FSGS), eGFR, and UPCR. Measures of fibrosis seen on the kidney biopsy (interstitial fibrosis and glomerular sclerosis) were thought to be potentially on the causal pathway and thus were not included in the models. Pearson’s correlation was used to assess the relationship between TNF score, biomarker tissue mRNA expression and urinary biomarker concentration. Linear regression models were fit to assess the association of urinary biomarkers and clinical features with TNF score and calculate a predicted score. Correlation between predicted and observed TNF activation score was assessed using Pearson’s correlation. Analyses were performed using STATA, v12.1 (College Station, TX) with two-sided tests of hypotheses and p-value <0.05 as the criterion for statistical significance.

### Single nuclear RNA-seq data processing

Nuclei were prepared from kidney biopsies tissue preparations stored in RNAlater using protocols developed and adapted from the Kidney Precision Medicine Project^31^. Analysis preparations were processed using 10x Genomics single cell sequencer. Analyses were performed on the output data files from CellRanger using the Seurat R package (version 3.2 and 4.0; https://cran.r-project.org/web/packages/Seurat/index.html). To limit low quality nuclei and/or multiplets, we set minimum and maximum cutoffs for gene counts per nuclei to 500 and 5000 genes, respectively and limited the analysis to nuclei with a mitochondrial gene content less than 10%. Nuclei were merged into a Seurat object using integrate function for downstream analyses. Nuclear cluster annotation was determined by first defining enriched genes in each cell cluster, and comparison of cluster selective gene profiles against previously identified cell marker gene sets from human kidney samples^30, 31^. Data submitted to GEO.

### Cell & kidney organoid culture

Culture of UM77-2 human embryonic stem cells (hESC), and generation of kidney organoids were performed as previously described^32^. Organoids were treated with TNFα (R&D Systems, Cat# 10291-TA) resuspended in PBS (Gibco #14190144) on D23 of cell culture at the indicated concentration. At indicated time points, organoid supernatant was removed and either stored temporarily at 4°C or frozen at −80°C. For RNA extraction, organoid Organoid wells were then rinsed twice with ice-cold PBS, scraped, and the cells pelleted. Cell pellets were lysed in TRIzol (Invitrogen, Cat#15596026) and Direct-zol RNA Miniprep Plus columns (Zymo Research, cat# R2072) with on-column DNase treatment (Zymo Research, cat# E1011-A). Organoid culture supernatants and cell lysates were diluted 150-fold to measure MCP-1 and 10-fold to measure TIMP-1 using the same ELISA protocol as the urine biomarker profiling described above.

### qRT-PCR

RNA quantity and quality was assessed by Nanodrop (ThermoFisher) via 260/280 ranging 1.91-2.04. One μg of total RNA was then reverse-transcribed into cDNA using SuperScriptFirst-Strand kit (Invitrogen, Cat# 11904-018) per manufacturer’s protocol. Quantitative real-time PCR was performed on a QuantStudio™ 7 Flex Real-Time PCR System (ThermoFisher) with sample analysis performed in triplicate using TaqMan Fast Universal PCR Master Mix (2X) (Applied Biosystems, Cat# 4352042) with the following Taq-Man Assay Reagents (Thermo Fisher): CCL2 (Hs00234140_m1), TIMP1(Hs01092512_g1), and GAPDH (Hs03929097_g1). The ΔΔCq method was applied to calculate the relative quantity (RQ, or fold change) of target genes after normalization to GAPDH. Graphs were plotted using GraphPad Prism software.

## Results

### Unbiased Consensus Clustering of Gene Expression Profiles to Identify Molecular Subgroups

Transcriptomes from NEPTUNE participants clustered into three groups (n=85, 76 and 59, respectively), with one cluster (T3) demonstrating the highest cluster stability (Figure 2A). The delta-K revealed that the 3 cluster solution was optimal across clustering approaches (Supplementary figure S1A). To validate the molecular profiles identified in Cluster 3, consensus clustering was applied to the tubulointerstitial transcriptome data from two independent cohorts. ERCB (N=30) and H3 Africa (N=35) patients also formed three distinct clusters (Figure 2B and C) with high cluster stability (T3). Glomerular clustering also identified 3 clusters (Supplementary Figure S2A, B and C) with a transcriptional signature largely shared with the tubulointerstitium (Figure 2D). To determine whether these high stability clusters are driven by a common molecular profile, differential expression was performed between cluster 3 and the other two clusters in each cohort and a robust directionally conserved molecular signal was seen (correlation of fold change 0.94, p<0.001 for NEPTUNE vs. ERCB and 0.93, p<0.001 for NEPTUNE vs. H3 Figure 2E and F). NEPTUNE participants in cluster 3 were older, and had a lower eGFR, greater interstitial fibrosis and higher UPCR at biopsy (Table 1, supplementary figure S3). In ERCB and H3Africa, participants in cluster 3 also had lower eGFR and were of older age. Although the high stability cluster T3 had a greater proportion with FSGS in all three cohorts, all three also had participants with MCD according to conventional morphologic criteria (Figure 2G, supplementary figure S1 D and E). Regarding clinical outcomes, in an unadjusted survival model, NEPTUNE Cluster 3 had a more aggressive phenotype, with a greater hazard of the composite of ESKD or 40% decline in eGFR [unadjusted HR 5.23 (95% CI 1.9, 14.5) for cluster 3 vs. 1, p<0.001 for overall differences in curves, Figure 2H] and fewer complete proteinuria remission events [unadjusted HR 0.73 (95% CI 0.43, 1.26) for cluster 3 vs. 1, p=0.068 for overall difference in curves, Figure 2I]

**Figure 2:**
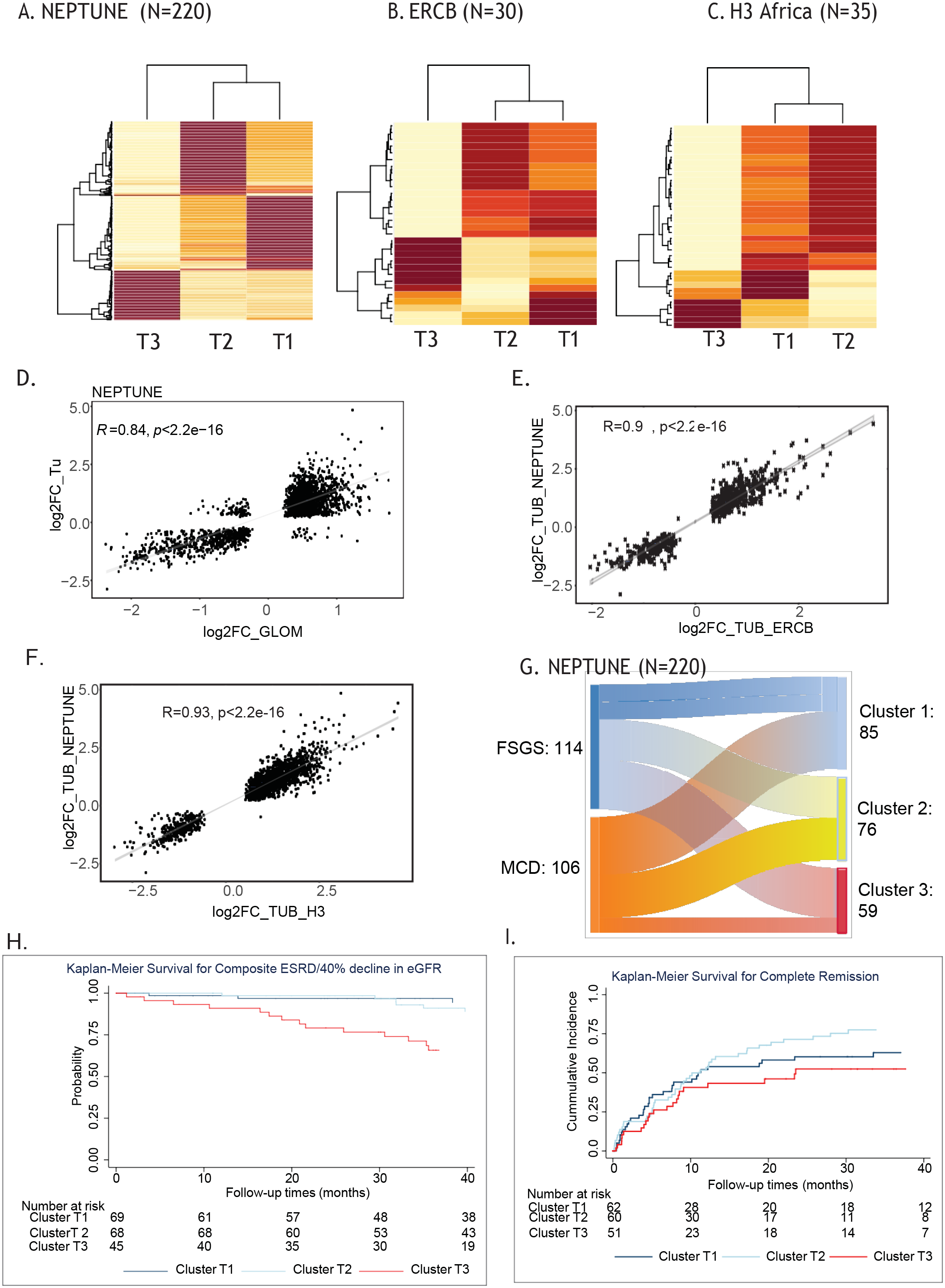
Kidney transcriptomic cluster membership and unadjusted Kaplan Meier curves. Consensus clustering was used to identify optimal cluster membership from TI transcriptomic profiles with 3 clusters in a cluster matrix from (A) NEPTUNE, (B) ERCB, and (C) H3 Africa cohorts. The values range from 0 (pale yellow, samples do not cluster together) to 1 (brown, samples demonstrate high affinity and cluster together). Scatter plots of significant fold change differences of genes differentially expressed in tubulointerstitial and glomerular compartments in NEPTUNE (D); cluster T3 compared to T2 and T1 from NEPTUNE (y-axis) compared to fold change differences of genes differentially expressed in cluster T3 compared to T1 and T2 from (E) ERCB (x-axis) and (F) H3 Africa (x-axis). (G) Alluvial plot demonstrating the relationship between diagnosis and cluster membership in the NEPTUNE cohort. Unadjusted Kaplan Meier survival curves by NEPTUNE cluster for (H) complete remission and (I) composite endpoint of 40% loss of EGFR or ESRD.

**Table 1.** Clinical characteristics of participants summarized by cluster identity and cohort

| NEPTUNE | ALL<br>(N = 220) | Cluster1<br>(N = 85) | Cluster 2<br>(N = 76) | Cluster 3<br>(N = 59) | P-value |
| --- | --- | --- | --- | --- | --- |
| AGE mean (sd) | 28.19 (21.65) | 22.27 (19.51) | 26.84 (22.62) | 38.47 (20.00) | P < 0.0001 |
| eGFR mean (sd) | 89.54 (46.80) | 102.06 (40.66) | 104.18 ( 48.86) | 52.26 (29.18) | P < 0.0001 |
| UPCR median (IQR) | 1.4 (0.3, 3.9) | 0.9 (0.2, 3.9) | 1.3 (0.2, 3.0) | 2.6 (1.0, 6.1) | P <0.0001 |
| % IF median (IQR) | 4 (0, 16) | 1.5 (0,5) | 3 (0, 8) | 20 (10, 55) | P <0.0001 |
| Disease duration prior to biopsy<br>(months) Median (IQR) | 4 (1, 25) | 5 (1, 37) | 3 (1, 13) | 2.5 (0, 34) | 0.3 |
| Female | 89 (40.45%) | 33 (38.82) | 30 (39.47%) | 26 (44.07%) | P = 0.8152 |
| On RAAS Blockade | 86 (39.09%) | 34 (40.00%) | 26 (34.21%) | 26 (44.07%) | P = 0.0005 |
| FSGS | 114 (51.82%) | 32 (37.65%) | 37 (48.68%) | 45 (76.27%) | P < 0.0001 |
| On IST | 101 (45.91%) | 49 (57.65%) | 37 (48.68%) | 15 (25.42%) | P = 0.0005 |
| ERCB | Cluster 1<br>(N = 18) | Cluster 2<br>(N = 4) | Cluster 3<br>(N = 8) | P-value |  |
| Age mean (sd) | 42.17 (18.97) | 29.36 (6.61) | 49.16 (18.28) | 0.63 |  |
| eGFR mean (sd) | 92.18 (34.51) | 119.01 (4.04) | 43.18 (30.96) | 0.02 |  |
| Female | 10 (55.56%) | 1 (25.00%) | 3 (37.50%) | 0.59 |  |
| H3 Africa | Cluster1<br>(N = 14) | Cluster 2<br>(N = 16) | Cluster3<br>(N = 5) | P-value |  |
| eGFR mean (sd) | 89.80 (42.58) | 109.78 (14.65) | 25.75 (11.73) | P = 0.0303 |  |
| UPCR mean (sd) | 1.78 (0.94) | 1.99 (2.19) | 4.68 (3.25) | P = 0.0486 |  |
| IF mean (sd) | 4.29 (5.84) | 2.19 (4.46) | 26.00 (24.08) | P = 0.0102 |  |
| FSGS: FSGS | 7 (50.00%) | 4 (25.00%) | 4 (80.00%) | P = 0.0681 |  |

### Biological and molecular relevance of cluster 3

Differential mRNA expression profiles from each cohort were used to elucidate the molecular functions associated with cluster 3. In NEPTUNE, there were 2721 transcripts in cluster 3 with a 1.5 fold-change and q<0.05, compared to clusters 1 and 2. This gene set was analyzed to assess enriched canonical pathways, to perform causal analysis to identify predicted upstream regulators controlling the transcriptional profile in cluster 3^33^, and to identify gene interaction networks in cluster 3.

The above analyses converged on TNF pathway activation. In signal transduction pathway over-representation (enrichment), the granulocyte adhesion and diapedesis signal transduction pathway had the highest enrichment score (-log(p)=21.4). Of the 180 genes in this pathway, 70 (38.9%) genes were found differentially regulated in cluster 3, including TNF (2.4-fold up-regulated in cluster 3, q<0.001), as a key immune response factor that induces pathway activation in endothelial cells (Figure 3A). Next, in a causal analysis of predicted upstream regulators, TNF was predicted as the top mediator activated in patients in cluster 3 (IPA network Z-score=13.2, enrichment p=2.09E-120). An expanded causal mechanistic network centered on downstream effects of predicted TNF activation (Figure 3B) explained 48% (1299/2721) of the differentially expressed genes between cluster 3 and clusters 1 and 2. Regulated transcripts included multiple transcription factors previously implicated in chronic kidney disease progression including NFκB (including NFκB 1 (p105/p50) and RELA (p65) subunits)^11, 22, 34^, and STAT1 and STAT3^35^. In a gene interaction network analysis, TNF was identified at the hub gene connecting the cluster 3 regulated gene set (Figure 3C). Mapping the upstream regulators across cohorts recapitulated the NEPTUNE signals in the ERCB and H3 Africa cohorts, with TNF identified as the top upstream regulator (Supplementary Table S1).

**Figure 3.**
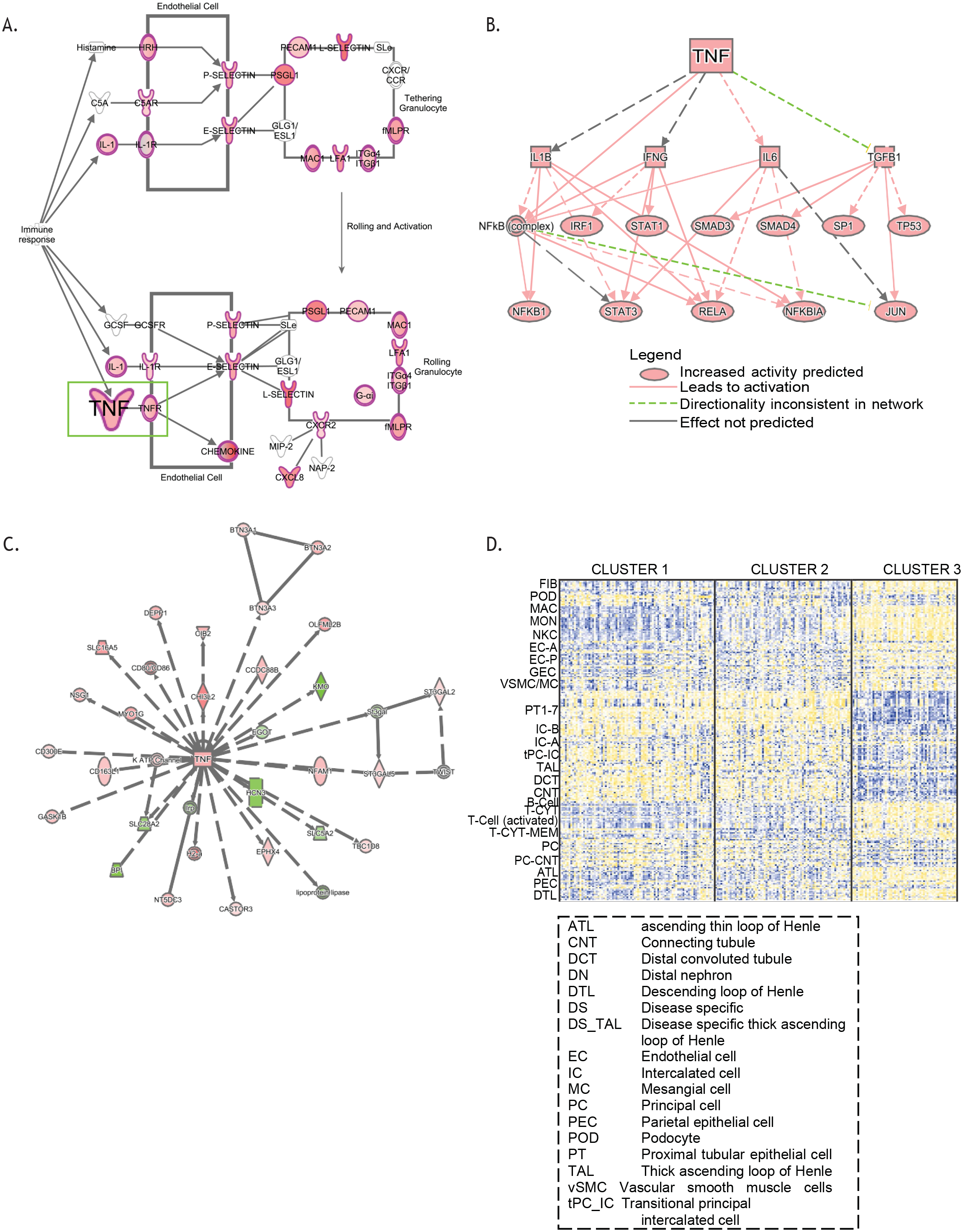
Molecular and functional context of cluster 3 expression profiles. Differential expression profiles from T3 compared to T1 and T2 in the NEPTUNE cohort were generated and enrichment analysis was performed using Ingenuity Pathways Analysis. (A) Granulocyte adhesion and diapedesis was the top enriched canonical pathway; a subset of the pathway is shown highlighting TNF as an input to the pathway. Genes highlighted in red were up-regulated in the differential expression profile. (B) A mechanistic network of predicted upstream regulators from the differential expression profile indicating TNF as an input. (C) TNF was identified in a gene interaction network (red indicates the gene was up-regulated in the differential expression profile, while green indicates down-regulation). (D) Cell selective gene expression markers were previously identified (Menon et al., 2020) and were intersected with voom transformed gene (row) normalized expression data (yellow indicates higher expression, blue indicates lower expression) to elucidate probable cell contribution to differential expression profiles.

Next, we aimed to determine the cellular source of the transcriptional signal associated with cluster 3 using recently published human kidney single cell data sets^30^ The top 10 genes selectively enriched in cell types from single cell RNAseq expression profiles from 24 human reference kidney tissues^30^ (Adult normal kidney dataset at nephrocell.miktmc.org) served as cell type specific markers. The marker set showed increased expression in cluster 3 in both kidney (fibroblasts, endothelial, parietal epithelial (PEC), ascending thin loop of Henle (ATL) and descending loop of Henle (DTL)) and immune cell lineages (macrophages, monocytes, NK cells, T-cells). The marker genes for proximal tubules, intercalated cells, thick ascending loop of Henle (TAL), distal convoluted tubule (DCT), connecting tubule (CNT), principal cells (PC), and PC-CNT showed lower steady state level in cluster 3 (Figure 3D). Taken together, these findings support TNF activation in transcriptional profiles from participants in cluster 3 and a molecular signature derived from resident kidney and immune cells.

### Patient-level TNF activation score and relationship to cluster information

Because multiple lines of evidence converged in TNF, we sought an approach to assess intra-kidney activation of TNF non-invasively for patient stratification. Using a rich knowledge base^36–38^, we extracted a set of 272 intra-renal transcript as a readout of TNF activity via its downstream regulated genes (supplementary table s2). The score was calculated for each patient biopsy profile^17, 39–41^, and evaluated across the cohorts (Figure 4A). Consistent with TNF activation accounting for a significant portion, the range of TNF score overlapped across the three cohorts (Figure 4A) with the highest scores in cluster 3. The TNF activation score was also calculated from glomerular samples and found to be strongly correlated with the tubulointerstitial TNF activation score in the two cohorts where matched gene expression samples were available (Figure 4B). A signature based on gene coefficients associated with TNF perturbation^42^ strongly correlated with our TNF activity score (R^2^>0.94, p<0.0001).

**Figure 4:**
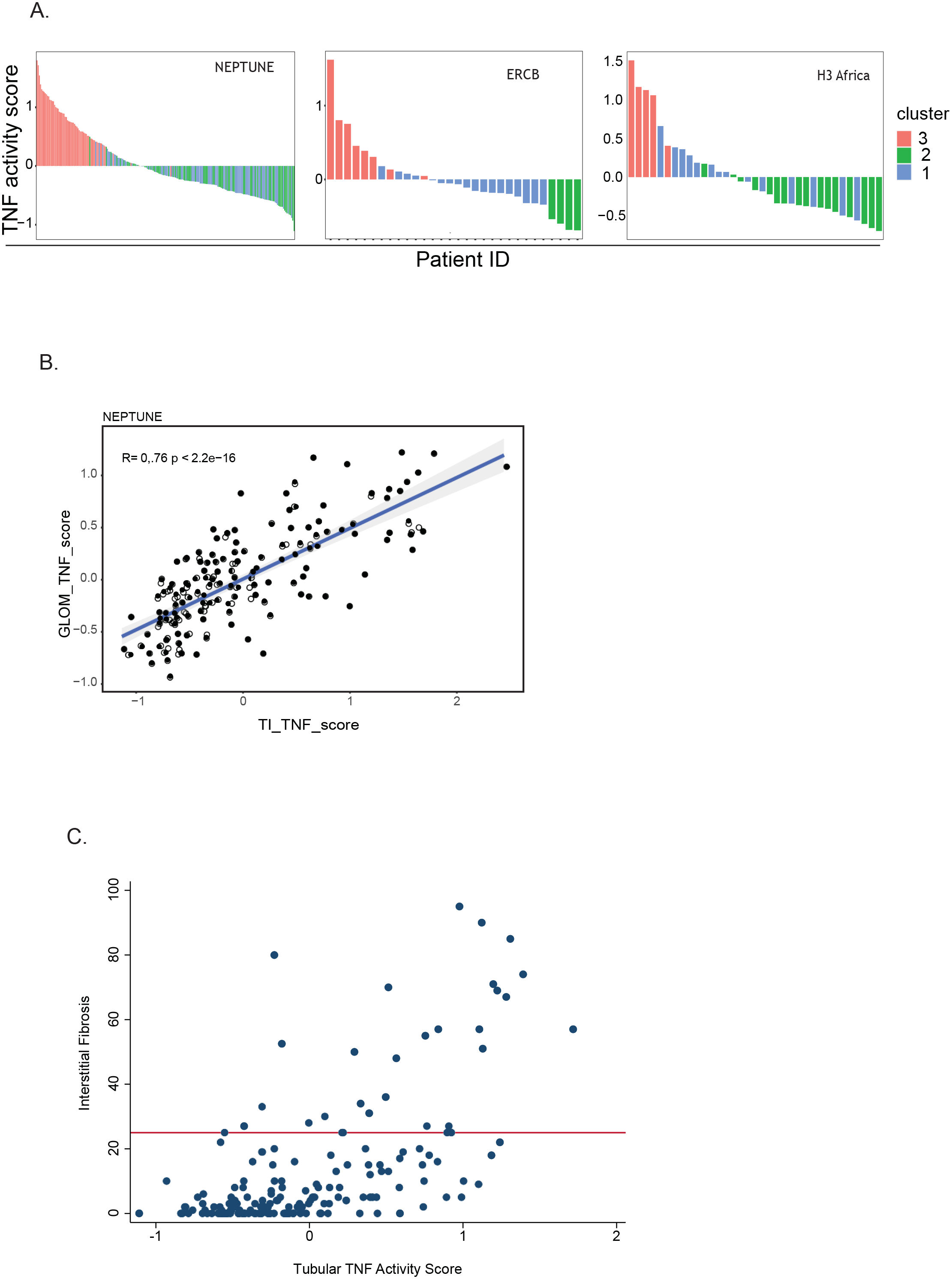
TNF activity scores across all profiled participants (A) from the indicated cohorts colored by cluster membership. (B) Time of biopsy log2 eGFR plotted by transcriptional cluster profile for patients in each cohort.

### Association of cluster 3 and TNF activation with interstitial fibrosis and clinical outcomes

The Spearman correlation of TNF activation scores with severity of interstitial fibrosis measured at the time of biopsy was significant (n=179, rho = 0.59, p<0.001, Figure 4C). Among the 148 participants with minimal interstitial fibrosis involving <25% of the kidney cortex, elevated TNF activation scores (TNF activation score >0) were observed in 47 (32%), indicating that the TNF activation score may be more sensitive to early signs of kidney damage than histopathological assessment.

To evaluate the extent to which the molecular information from the kidney tissue captured the variability in loss of eGFR over time observed in cluster 3 versus clusters 1 and 2, a survival model was fit separately with cluster membership (Model 1, Table 2) and TNF activation score (Model 2, Table 2) as primary predictors of interest. After adjustments for diagnosis (MCD vs. FSGS), baseline eGFR and UPCR, cluster 3 was associated with a higher hazard of reaching the composite outcome, HR 3.8, p=0.035. Cluster 2 was not significantly different from cluster 1. A 1 unit greater TNF activation score was associated with higher hazard of the composite outcome (unadjusted HR 2.6, p<0.001). After adjusting for diagnosis, the HR remained elevated (2.3, p=0.003). The association was attenuated after further adjustment for eGFR and UPCR (HR 1.7, p=0.12), suggesting that these factors are on the causal pathway of GFR decline.

**Table 2:** Unadjusted and adjusted Cox proportional hazards models for composite of ESKD and 40% decline in eGFR from baseline.

|  |  | Unadjusted Model |  | Adjusted for MCD/FSGS |  | Adjusted for MCD/FSGS, eGFR and UPCR |  |
| --- | --- | --- | --- | --- | --- | --- | --- |
|  | Predictor | HR<br>(95% CI) | p-value | HR<br>(95% CI) | p-value | HR<br>(95% CI) | p-value |
| **Model 1:**<br>**Cluster**<br>**Membership** | Cluster 1 | Ref |  | Ref. |  | Ref. |  |
|  | Cluster 2 | 1.7<br>(0.6, 5.1) | 0.34 | 1.6<br>(0.5, 4.8) | 0.40 | 2.3<br>(0.69, 7.57) | 0.18 |
|  | Cluster 3 | 5.2<br>(1.9, 14.5) | 0.001 | 4.5<br>(1.6, 12.9) | 0.005 | 3.80<br>(1.1, 13.1) | 0.035 |
| **Model 2: TNF**<br>**activation**<br>**score\*** | TNF<br>Activation<br>Score | 2.6<br>(1.5, 4.4) | 0.001 | 2.3<br>(1.3, 4.1) | 0.003 | 1.7<br>(0.9, 3.5) | 0.12 |

\*HR for increase in z-score by 1.
MCD, Minimal Change disease; FSGS, Focal segmental glomerulosclerosis; eGFR, estimated glomerular filtration rate (mL/min/1.73m<sup>2</sup>); UPCR, Urine protein to creatinine ratio (mg/mg)

### Identification of non-invasive biomarkers of TNF activation

Based on our previous studies assessing urine as a surrogate for intra-renal mRNA expression states^43^, we hypothesized that an intra-renal pathway activation signal may be reflected in participant urine profiles. In NEPTUNE, we cross-referenced genes in the TNF activity signature with the urine proteins profiled. Fourteen genes were identified (CCL2, CCL4, CCL5, CCL11, CXCL1, CXCL10, IL1RN, IL7, MMP2, MMP9, TIMP1, TIMP2, TNF) that had corresponding urinary proteins and are a subset of a TNF activation network (Figure 5A). Urine biomarkers with observed levels in the dynamic range in at least 75% of samples assayed, and those with a significant correlation (p<0.0001) between the associated intra-renal mRNA and the urine protein level were carried forward as representative of the intra-renal transcriptional state. Two genes, *CCL2* and *TIMP1* had intra-renal gene expression that were strongly correlated with urine biomarker level (r=0.58, p<0.0001 and r=0.50, p<0.0001, respectively). Urine biomarker profiles for MCP-1 (the protein encoded by *CCL2*) and TIMP-1 were also highly correlated with the TNF activation score (p<0.0001, r≥0.50 for both biomarkers, Figure 5B and 5C, respectively). Thus, these biomarkers were identified as potential non-invasive surrogates reflective of the intra-renal TNF activation score.

**Figure 5:**
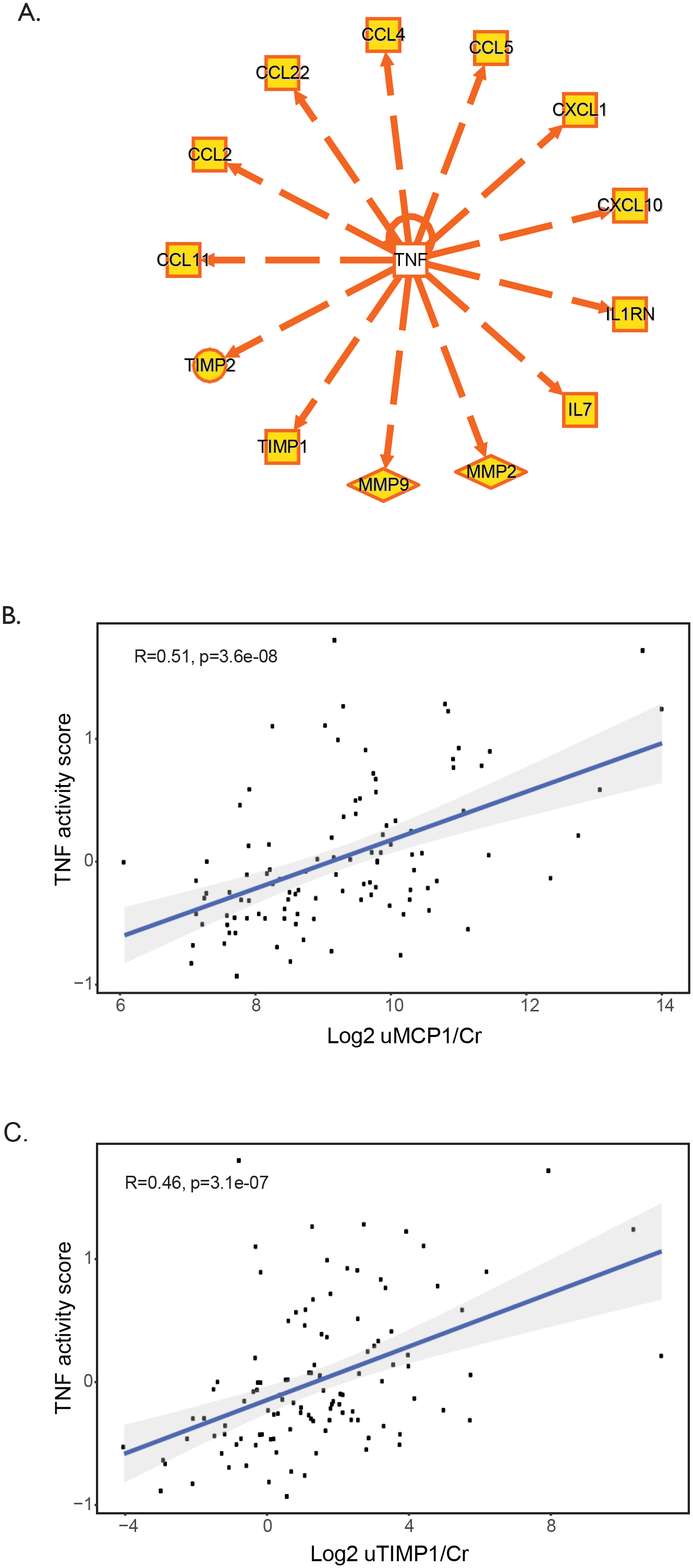
Biomarker selection for TNF activation. (A) Twelve genes up-regulated in cluster T3, were downstream of TNF activation through curated cause and effect relationships, and were present on the Luminex panel used to profile urine profile from NEPTUNE participants. TNF activity score plotted against (B) Log2 uMCP1/Cr and (C) Log2 uTIMP1/Cr.

### TNF effect on kidney organoids and resident kidney cell types

To further support the relationship of the non-invasive candidate biomarkers to intra-renal TNF activity, we tested their response to TNF stimulation in kidney organoids and defined their intra-kidney source using snRNAsequencing of NEPTUNE biopsies of patients with high and low TNF activation scores. First, TNF treatment of human kidney organoids resulted in up-regulation of *CCL2* and *TIMP1* mRNA expression (Figure 6A) followed by increased protein detection of the encoded proteins MCP-1 and TIMP-1 in the organoid supernatant (Figure 6B), demonstrating concordant induction of transcripts and proteins of the candidate biomarker. Then, to test the cellular source of the TNF pathway biomarker candidates, we performed snRNAseq on 10 NEPTUNE biopsies, five with high and five with moderate to low TNF activity scores in the tubulointerstitial gene expression profiles (Supplementary Table S3). Fifteen unique nuclear clusters were identified covering all major cell types of the kidney (Figure 6C) and evaluated for *CCL2* and *TIMP1* expression. Cell type specific gene expression pattern were found with consistently higher levels of both transcripts seen in the patients with elevated TNF scores. High expression were observed not only in immune cells, but also in intrinsic kidney cell clusters (Figure 6D). Thus, the TNF-responsive biomarkers reflect alterations in inflammatory and intrinsic kidney cell populations in patients with the TNF-associated signaling profile.

**Figure 6:**
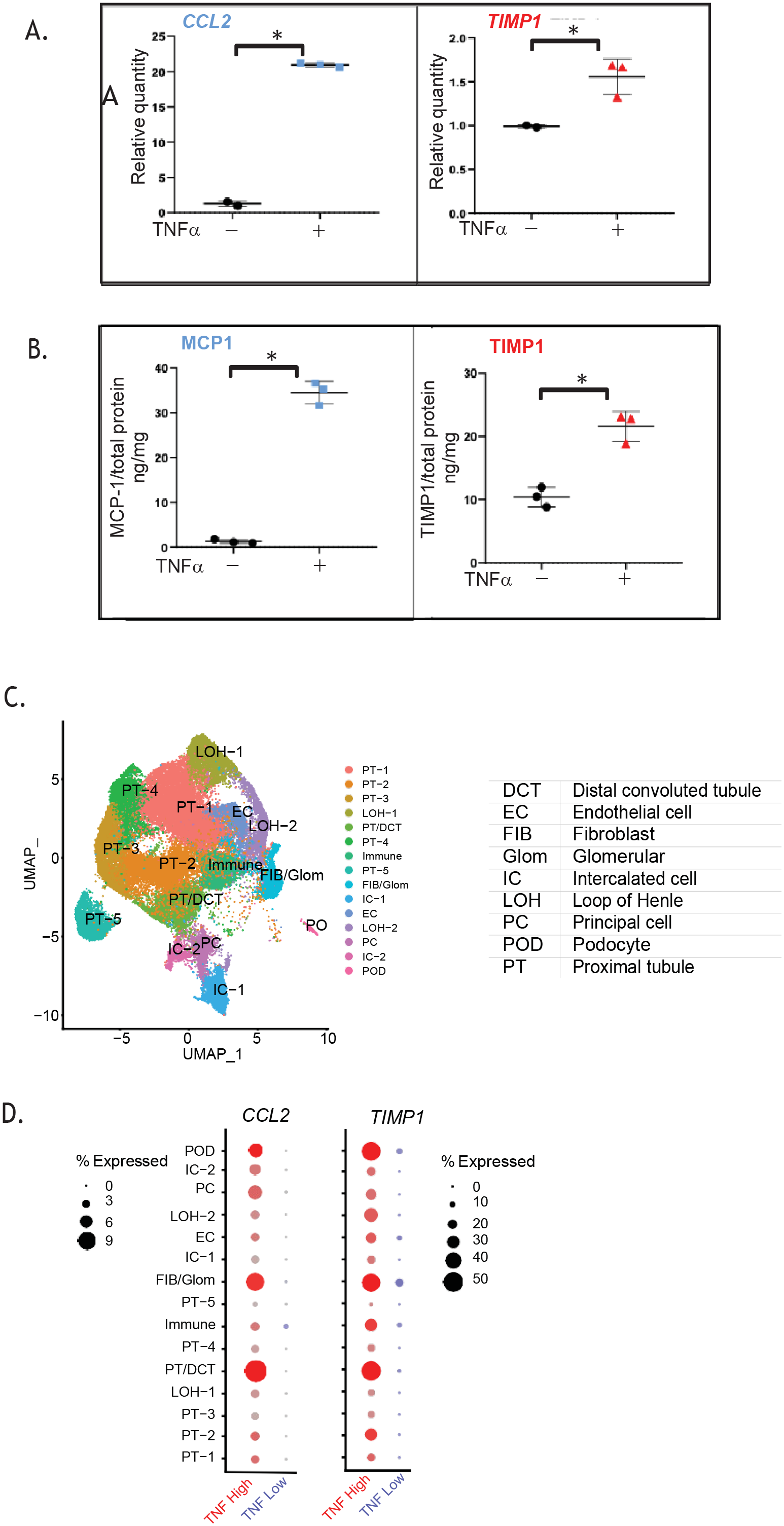
Tissue and cell source of TNF biomarkers. TNFα directly stimulates expression of selected biomarkers in human pluripotent stem cell derived-kidney organoids. Quantification of (A) *CCL2* (left) and *TIMP1* (right) transcript levels in kidney organoid cell lysates by qRT-PCR relative to control, and of (B) MCP-1 (left) and TIMP-1 (right) protein levels in kidney organoid culture supernatant by ELISA normalized to total protein, generated from the same samples following treatment with 5 ng/ml TNFα or vehicle control for 24h. Each data point was generated from a unique sample and represents the average of analysis in triplicate. Long bar, mean; short bar, 1 S.D.; *, p-value < 0.05 by Student’s t-test. Representative experiment (1 of 4 independent) shown. (C) UMAP plot of snRNAseq profiles from TI of selected NEPTUNE participants found to have elevated TNF activity scores (TNF high) and low to moderate TNF activity scores (TNF low) and (D) Single nuclear cluster expression of *CCL2* and *TIMP1* by TNF activity status.

### Predictive ability of biomarkers

NEPTUNE participants diagnosed with MCD or FSGS and TNF activation scores also had urinary cytokine measurements within 45 days post-biopsy (N=90). Using a combination of urine biomarkers (MCP-1 and TIMP-1), eGFR and UPCR, a predicted TNF score was calculated and highly correlated with the transcriptionally derived intra-renal TNF activation score (r=0.61, p<0.001), Figure 7. Thus, a biomarker with MCP-1 and TIMP-1 coupled with routine clinical information can predict intra-renal TNF activation profiles in patients with FSGS and MCD.

**Figure 7:**
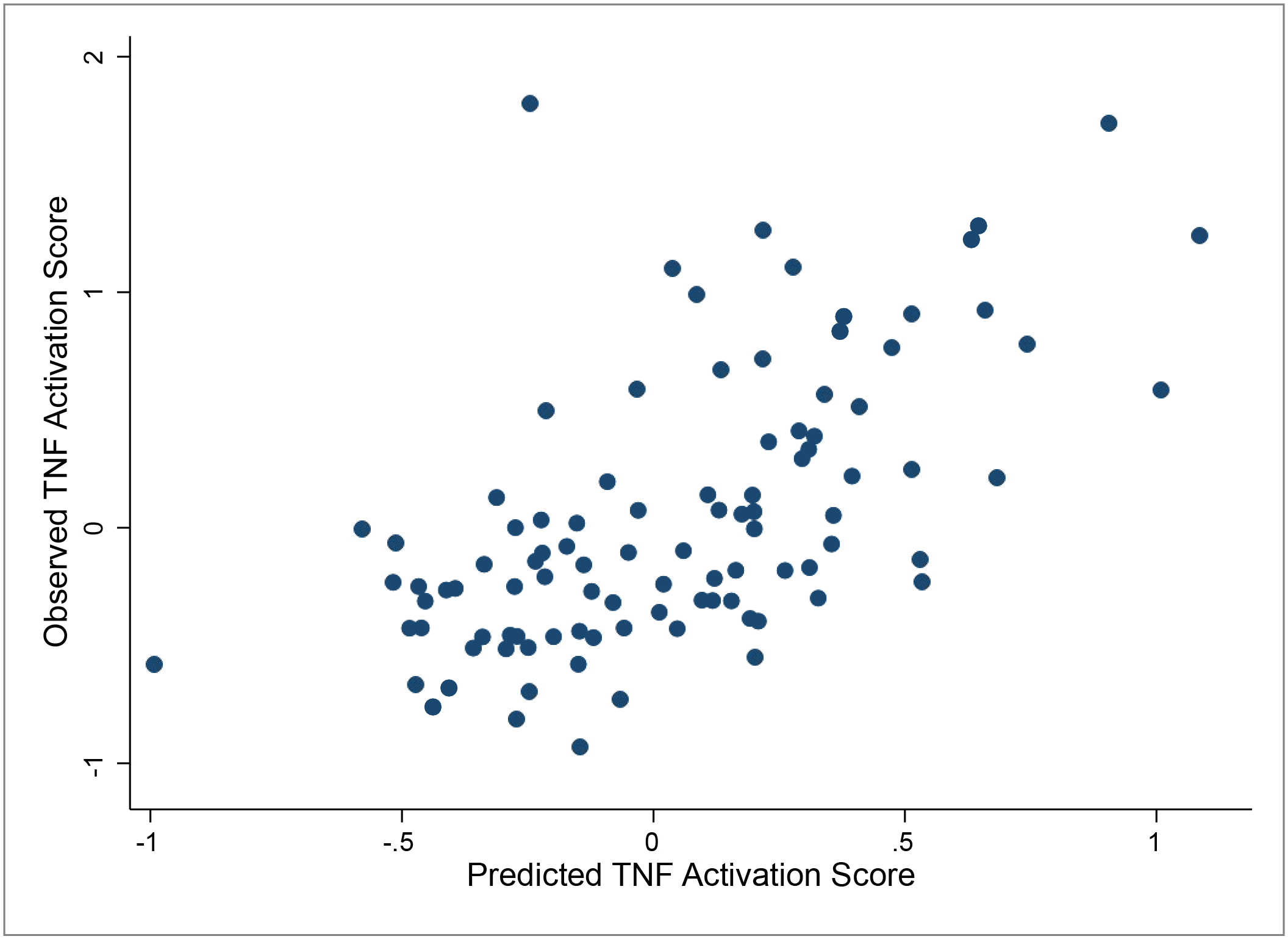
Correlation of observed TNF activation score with a predicted score based on urinary biomarkers and clinical features. Linear regression models were used to generate predicted tissue TNF activation scores based on eGFR, UPCR, urinary TIMP1 and urinary MCP1. Correlation was 0.61, p-value <0.001.

## Discussion

This study aimed to address the heterogeneity in presentation, clinical course, and response to treatment in patients with glomerular diseases with the goal of moving the field towards precision medicine^1^. Our objective was to define biologically and clinically relevant patient subgroups with specific activation pathways and corresponding non-invasive biomarkers.

The study leveraged kidney tissue transcriptomics to identify subgroups of patients defined by a shared molecular profile and poor clinical outcomes across three geographically diverse cohorts. The molecular classification was independently associated with clinical outcome, even after adjusting for histopathology diagnosis and laboratory measures, suggesting that it captures prognostic information not contained in current evaluation protocols.

The molecular profile of this group was evaluated for underlying cellular and biological processes and found to center on TNF activation, a cytokine linked to a range of diseases ^44–47^. TNF is produced by infiltrating immune cells as well as resident kidney cells, including podocytes^48^ and mesangial cells^49^. In isolated rat glomeruli, TNFα administration increased albumin permeability^50^. In Buffalo/Mna rats that spontaneously develop a form of FSGS, increased kidney expression of TNF precedes the onset of proteinuria^51^. TNF synthesized in the kidney induces podocyte damage through cholesterol-dependent apoptosis irrespective of serum levels. In vitro exposure of podocytes to serum obtained from patients with primary FSGS activates inflammatory pathways including TNF. Moreover, the degree of TNF activation correlates with clinical and histopathological indices of disease severity^52^. In addition to glomerular signaling, kidney tubular epithelial cells express TNF Receptor 2 (TNFR2), and manifest inflammatory features when exposed to TNF^53^. Finally, mice with macrophage-specific TNF deletion have recently been shown to have less interstitial fibrosis^54^. This indicates that TNF impacts all kidney tissue compartments and cell types. Glomerular *TNF* mRNA expression was negatively correlated with eGFR in NEPTUNE participants with FSGS^48^. In humans, TNF levels from cultured peripheral blood mononuclear cells were higher in children with active nephrotic syndrome, compared to those in remission and controls^47^. A subset of patients with FSGS were shown to have TNF pathway activation in glomeruli ^52–55^. These findings are supported by the cell-type specific deconvolution approach on the bulk transcriptomic data in the current study which showed that transcriptional signals characterizing the high TNF cluster were derived from infiltrating immune cells and nephron lineage cells including endothelial cells. This was confirmed in the snRNAseq data where participants with high TNF scores showed increased expression of *CCL2* and *TIMP1* in multiple cell types.

In our study, TNF activation in the kidney tissue was sufficient to capture the association with poor clinical outcome. Two candidate non-invasive biomarkers of TNF activity, combined with existing clinical measures, were accurate predictors of intra-renal TNF activation. Although TNF activation was associated with the degree of fibrosis in some kidney biopsies, many patients without significant scarring had elevated TNF activation, demonstrating pathway activation early in the disease course. The two identified biomarkers were MCP-1 a marker of active inflammation^56^ and TIMP-1 which is associated with tissue remodeling and scarring^57^.

Case reports and small studies have reported that anti-TNF therapy may be effective in a subset of nephrotic syndrome patients, but none considered intra-renal activation of the pathway prior to therapy as a method for stratifying patients^58–60^. For example, the FONT trial (Novel Therapies for Resistant FSGS) tested the TNF inhibitor adalimumab in patients with multi-drug resistant FSGS using an unstratified approach^61, 62^. Of the 17 patients treated in the combined phase I and phase II studies, 4 had ≥50% reduction in proteinuria with 2 patients achieving dramatic improvements, from UPCR of 17 to 0.6 mg/mg in one and from 3.6 to 0.6 mg/mg in a second. Although the study was considered unsuccessful in demonstrating efficacy of anti-TNF therapy for all FSGS patients, a response in any patient with this severe phenotype is notable and consistent with significant underlying study sample heterogeneity.

The unpredictable outcome in FONT is consistent with other trials, where despite overall negative results, small subsets of patients appeared to respond to the tested therapy. Treatment with adrenocorticotropic hormone (ACTH) gel achieved a complete or partial remission in 7 out of 24 FSGS patients (29%)^63^. Although a case series suggested that abatacept would be a useful agent for the treatment of FSGS^64^, a randomized clinical trial that included all patients with treatment-resistant FSGS was terminated prematurely because of lack of efficacy. These findings highlight the need for a precision medicine approach to match the disease mechanism with the targeted treatments being tested.

Clinical trials could incorporate markers - serum, urine, or genetic-suggestive of activation of a relevant injury pathway and higher risk of disease progression. This approach would promote an alignment of the molecular profile of the trial participants with the mechanism of action of the investigational agent. Molecular categorization could be combined with consensus clustering based on clinical and laboratory data, as outlined here for nephrotic syndrome and recently applied to the CRIC cohort^65^ for precise delineation of patient prognosis and optimization of therapy.

Several limitations of our approach are acknowledged. The clustering was done using the tubulointerstitial compartment as opposed to the glomerular compartment. However, tubulointerstitial damage and fibrosis has been shown to be one of the strongest predictors of clinical outcome in the NEPTUNE cohort and treatment response^17^ across conventional disease classifications, and we see a strong correlation of cluster-associated signatures across tissue compartments. In addition, our molecular clusters associated strongly with eGFR outcomes. Additionally, the TNF activation signal in the matching glomerular compartment strongly correlated with tubulointerstitial TNF activation, likely capturing similar activation in both compartments.

For some patients, high TNF activation may represent disease that is too advanced to be responsive to the proposed therapy. However, one third of patients showed low interstitial fibrosis but high TNF scores, indicating that TNF activation can be present without fibrosis. Thus, TNF activation may precede disease progression^51^. While genetic information is expanding our understanding of nephrotic syndrome, in this cohort of incident nephrotic syndrome patients, a low frequency of mutation rates in 21 monogenic nephrotic syndrome genes was seen in NEPTUNE^66^, preventing us from finding an association of TNF with monogenic disease. Longitudinal studies are needed to validate the use of TNF activation scores as a target engagement biomarker during the treatment of FSGS and other glomerular diseases. Finally, revisiting the FONT trial coupled with a patient stratification approach to limit trial eligibility to patients with high predicted pathway activity will enable assessment of whether the pathway activity score is responsive the TNF inhibition. A Phase IIa trial with this design has been initiated (clinicaltrials.gov/NCT04009668).

In conclusion, this study implements a precision medicine strategy, identifies a molecularly defined subgroup of patients with poor clinical outcomes and a targetable pathway, TNF, as a potential key driver of disease progression. Further, non-invasive markers, validated in an organoid model system, are available to identify the subgroup with TNF activation, an approach currently tested in an interventional trial. The concept of mechanistic disease classification system developed here for FSGS/MCD and the TNF pathway represents a first step for a comprehensive map projecting glomerular diseases into a landscape of targetable pathways and a move towards precision medicine for glomerular diseases.

## Data Availability

Transcriptional profiles of biopsies from patients with MCD and FSGS in the ERCB have been deposited to GEO and are part of accession numbers GSE104954 (tubulointerstitium) and GSE104948 (glomeruli).

https://www.nephroseq.org/resource/login.html

## Author Contributions

All co-authors have contributed to the manuscript. LHM, SE, MK, KRT, CS, MS, PHN, AA, JCL, CCD, DA, KLG, JRS, KEM, HNR, SMB, RAL, JBH, DSG, JLH conceptualized this study; LHM, SE, VB,WJ, SB, JLH, J-JL, FMA, MS, VN, PJM contributed to data curation; MHL, SE, FMA, PJM, WJ, JLH did the formal analysis; MHL, SE, MK, VB, DA, JCL, JRS, AF, LBH, RAL, LB, DSG, AOO were involved in funding acquisition; MHL, SE, MK, PJM, KRT, VB, KVL, LAB, SGA, ADA, TS, CW, FF, CLT, DCC, WJ, DA, KLG, BG, KEM, LBH, SMB, RAL, JBH, MAA, J-JL, investigated the findings; FMA, SM, PJM, BG, RAL, JBH, VN, SE, JLH developed the methodology; MHL, SE, KRT, VB, FE, KKS, TS, FF, SMV, DA, KLG, KEM were involved in project administration; KVL, LAG, SGA, ADA, AA, LHM, JCL, FF, SMV, CLT, DA, KLG, BG, AF, KEM, HNR, LBH, RAL, JBH, MAA, J-JL, MK provided resources; SE, FMA, PJM worked on software used in the study; SE, FE, FMA, PJM performed bioinformatic processing and analyses in the study; LHM, SE, KRT, VB, ADA, AA, JCL, VKD, KLG, AF, KEM, HNR, BLH, RAL, MK, JLH supervised the study; VB, ADA and WJ worked on study validation NLW, VVW, ECT, JES performed kidney organoid experiments; BG, RM, PJM, EAO coordinated processing and analysis of single nuclear RNAseq samples; LHM, SE, VB, CS, FMA, ADA, PJM, KEM, LBH, RAL, helped with visualization; LHM, SE, KRT, VB, RAL wrote the original draft; all authors reviewed and provided valuable feedback on the manuscript.

## Acknowledgement

We thank Dr. Lalita Subramanian for help with writing, editing and formatting this manuscript.The Nephrotic Syndrome Study Network Consortium (NEPTUNE), U54-DK-083912, is a part of the National Institutes of Health (NIH) Rare Disease Clinical Research Network (RDCRN), supported through collaboration between the Office of Rare Diseases Research, National Center for Advancing Translational Sciences and the National Institute of Diabetes, Digestive, and Kidney Diseases. Additional funding and/or programmatic support for this project has also been provided by the Else Kröner-Fresenius Foundation (ERCB), University of Michigan, the NephCure Kidney International and the Halpin Foundation, and the Applied Systems Biology Core at the University of Michigan George M. O’Brien Kidney Translational Core Center (2P30-DK-08194). Dr Mariani is supported through funding from NIH/NIDDK, K08 DK115891-01. We acknowledge the role of the H3Africa Consortium in making this research possible though the sharing of data and knowledge. The National Institutes of Health (USA) and Wellcome Trust (UK) have provided the core funding for the H3Africa Consortium and more information is available at https://h3africa.org/. This research was supported by the following grants from NIH/NHGRI/NIDDK: H3Africa Kidney Disease Study (U54 HG006939), H3Africa Kidney Disease Cohort Study (U01 DK107131), H3Africa Kidney Disease Collaborative Centers (9U54 DK116913). The views expressed in this paper do not represent the views of the H3Africa Consortium or their funders. ERCB, NEPTUNE and H3 Africa contributing members are listed in supplemental acknowledgement.

## List of Supplementary Materials

1. Supplementary acknowledgements

a. ERCB
b. NEPTUNE
c. H3 Africa
2. Supplementary Figure S1. Cluster dendrogram and assignment of MCD and FSGS participants based on kidney biopsy tubulointerstitial gene expression data in NEPTUNE, ERCB and H3 Africa
3. Supplementary Figure S2. Comparison of cluster assignment and clinical measures in NEPTUNE and ERCB using glomerular compartment expression data
4. Supplementary Figure S3. Comparison of clinical factors and cluster assignment across cohorts
5. Supplementary Table S1. Predicted Upstream Regulators based on DEG profiles of patients in cluster 3 relative to clusters 1 and 2.
6. Supplementary Table S2. Genes and gene products activated by TNF used to generate the TNF activation score.
7. Supplementary Table S3. Clinical characteristics of NEPTUNE participants with biopsies used for snRNAseq

