## Supplementary materials for "Multidimensional Data Integration Identifies Tumor Necrosis Factor Activation in Nephrotic Syndrome: A Model for Precision Nephrology"

**List of Supplementary Materials**

1. Supplementary acknowledgements
    a. ERCB
    b. NEPTUNE
    c. H3 Africa
2. Supplementary Figure S1. Cluster dendrogram and assignment of MCD and FSGS participants based on kidney biopsy tubulointerstitial gene expression data in NEPTUNE, ERCB and H3 Africa
3. Supplementary Figure S2. Comparison of cluster assignment and clinical measures in NEPTUNE and ERCB using glomerular compartment expression data
4. Supplementary Figure S3. Comparison of clinical factors and cluster assignment across cohorts
5. Supplementary Table S1. Predicted Upstream Regulators based on DEG profiles of patients in cluster 3 relative to clusters 1 and 2.
6. Supplementary Table S2. Genes and gene products activated by TNF used to generate the TNF activation score.
7. Supplementary Table S3. Clinical characteristics of NEPTUNE participants with biopsies used for snRNAseq

**ERCB members at the time of the study**: Clemens David Cohen, Holger Schmid, Michael Fischereder, Lutz Weber, Matthias Kretzler, Detlef Schlöndorff, Munich/Zurich/AnnArbor/New York; Jean Daniel. Sraer, Pierre Ronco, Paris; Maria Pia Rastaldi, Giuseppe D'Amico, Milano; Peter Doran, Hugh Brady, Dublin; Detlev Mönks, Christoph Wanner, Würzburg; Andrew Rees, Aberdeen and Vienna; Frank Strutz, Gerhard Anton Müller, Göttingen; Peter Mertens, Jürgen Floege, Aachen; Norbert Braun, Teut Risler, Tübingen; Loreto Gesualdo, Francesco Paolo Schena, Bari; Gunter Wolf, Jena; Rainer Oberbauer, Dontscho Kerjaschki, Vienna; Bernhard Banas, Bernhard Krämer, Regensburg; Moin Saleem, Bristol; Rudolf Wüthrich, Zurich; Walter Samtleben, Munich; Harm Peters, Hans-Hellmut Neumayer, Berlin; Mohamed Daha, Leiden; Katrin Ivens, Bernd Grabensee, Düsseldorf; Francisco Mampaso(†), Madrid; Jun Oh, Franz Schaefer, Martin Zeier, Hermann-Joseph Gröne, Heidelberg; Peter Gross, Dresden; Giancarlo Tonolo; Sassari; Vladimir Tesar, Prague; Harald Rupprecht, Bayreuth; Hermann Pavenstädt, Münster; Hans-Peter Marti, Bern; Peter Mertens, Magdeburg, Jens Gerth, Zwickau.

**Members of the Nephrotic Syndrome Study Network (NEPTUNE) at the time of the study**

NEPTUNE Enrolling Centers
*Case Western Reserve University, Cleveland, OH*: J Sedor[*], K Dell**, M Schachere[#], J Negrey
*Children's Hospital, Los Angeles, CA*: K Lemley[*], L Whitted[#]
*Children's Mercy Hospital, Kansas City, MO*: T Srivastava[*], C Haney[#]
*Cohen Children's Hospital, New Hyde Park, NY:* C Sethna[*], K Grammatikopoulos[#], R Odusayana
*Columbia University, New York, NY:* G Appel[*], M Toledo[#]
*Emory University, Atlanta, GA:* L Greenbaum[*], C Wang**, B Lee[#]
*Harbor-University of California Los Angeles Medical Center:* S Adler[*], C Nast[*‡], J La Page[#]
*John H. Stroger Jr. Hospital of Cook County, Chicago, IL:* A Athavale[*]
*Johns Hopkins Medicine, Baltimore, MD:* M. Atkinson, A Neu[*], S Boynton[#]
*Mayo Clinic, Rochester, MN:* F Fervenza[*], M Hogan**, J Lieske[*], V Chernitskiy[#]
*Montefiore Medical Center, Bronx, NY:* F Kaskel[*], N Kumar[*], P Flynn[#]
*NIDDK Intramural, Bethesda MD:* J Kopp[*], E Castro-Rubio[#], J Blake[#]
*New York University Medical Center, New York, NY:* H Trachtman[*], O Zhdanova**, F Modersitzki[#], S Vento[#]
*Stanford University, Stanford, CA:* R Lafayette[*], M O'Shaughnessy**, K Mehta[#]
*Temple University, Philadelphia, PA:* C Gadegbeku[*], D Johnstone**, S Quinn-Boyle
*University Health Network Toronto:* D Cattran[*], M Hladunewich**, H Reich**, P Ling[#], M Romano[#]
*University of Miami, Miami, FL:* A Fornoni[*], L Barisoni[*], C Bidot[#]
*University of Michigan, Ann Arbor, MI:* M Kretzler[*], D Gipson*, A Williams[#], R Pitter[#]
*University of North Carolina, Chapel Hill, NC:* V Derebail[*], K Gibson[*], S Grubbs[#], A Froment[#]
*University of Pennsylvania, Philadelphia, PA:* L Holzman[*], K Meyers**, K Kallem[#], A Swensen[#]
*University of Texas Southwestern, Dallas, TX:* K Sambandam[*], E Brown**, Z Wang[#]
*University of Washington, Seattle, WA:* A Jefferson[*], S Hingorani**, K Tuttle**[§], L Curtin[#], S Dismuke[#], A Cooper[#§]
*Wake Forest University, Winston-Salem, NC:* B Freedman[*], JJ Lin**, S Gray[#]
*Data Analysis and Coordinating Center*: M Kretzler, L Barisoni, C Gadegbeku, B Gillespie, D Gipson, L Holzman, L Mariani, M Sampson, P Song, J Troost, J Zee, E Herreshoff, S Li, C Lienczewski, T Mainieri, M Wladkowski, A Williams, D Zinsser
*National Institute of Diabetes and Digestive and Kidney Diseases (NIDDK) Program Office: K Abbott, C Roy*
*The National Center for Advancing Translational Sciences (NCATS) Program Office: T Urv, PJ Brooks*

**Principal Investigator; **Co-investigator; #Study Coordinator*
*‡Cedars-Sinai Medical Center, Los Angeles, CA*
*§Providence Medical Research Center, Spokane, WA*

**Members of the Human Heredity and Health in Africa Kidney Disease Research Network cohort (H3Africa)**

Samuel Ajayi[1], Yemi Raji[1], Timothy Olanrewaju[2], Charlotte Osafo[3], Ifeoma Ulasi[4], Adanze Asinobi[1], Cheryl A. Winkler[5], David Burke[6], Fatiu Arogundade[7], Ivy Ekem[8], Jacob Plange-Rhule[9**], Manmak Mamven[10], Michael Mate-kole[3], Olukemi Amodu[1], Richard Cooper[11], Sampson Antwi[9], Adebowale Adeyemo[12], Titilayo Ilori[13], Victoria Adabayeri[3], Alexander Nyarko[14], Anita Ghansah[14], Ernestine Kubi Amos-Abanyie[14], Priscilla Abena Akyaw[14], Babatunde L. Salako[1], Rulan S. Parekh[15], Bamidele Tayo[11], Rasheed Gbadegesin[16], Michael Boehnke[6] , Robert Lyons[6] , Frank (Chip) Brosius[6] , Daniel Clauw[6], Chijioke Adindu[2], Clement Bewaji[2], Elliot Koranteng Tannor[9], Perditer Okyere[9], Chuba Ijoma[4], Nicki Tiffin[17], Junaid Gamiedien[17], Friedhelm Hildebrandt[18], Charles Odenigbo[19], Nonyelun Jisieike-Onuigbo[19], Ifeoma Modebe[19], Aliyu Abdu[20], Patience Obiagwu[20], Ogochukwu Okoye[21], Adaobi Solarin[22], Toyin Amira[23], Christopher Esezobor[23], Muhammad Makusidi[24], Santosh Saraf[25], Victor Gordeuk[25], Gloria Ashuntangtang[26], Georgette Guenkam[26], Folefack Kazi[26], Olanrewaju Adedoyin[2], Mignon McCullough[27], Peter Nourse[27], Uche Okafor[4], Emmanuel Anigilaje[10], Patrick Ikpebe[22], Tola Odetunde[4], Ngozi Mbanefo[4], Wasiu Olowu[7], Paulina Tindana[3], Olubenga Awobusuyi[22], Olugbenga Ogedegbe[28], Opeyemi Olabisi[29], Karl Skorecki[30], Ademola Adebowale[1], Matthias Kretzler[6], Jeffrey Hodgin[6], Dwomoa Adu[3], Akinlolu Ojo[31], Vincent Boima[3]

**Affiliated Institutions**

1. Department of Medicine, Pediatrics and Institute of Child Health, University of Ibadan, Ibadan, Nigeria
2. University of Ilorin, Ilorin, Nigeria
3. University of Ghana Medical School, Accra, Ghana
4. University of Nigeria, Enugu State, Nigeria
5. Basic Research Laboratory, Frederick National Laboratory for Cancer Research, National Cancer Institute, Frederick, MD, USA
6. Departments of Human Genetics, Internal Medicine and Pathology, University of Michigan, Ann Arbor, MI, USA
7. Obafemi Awolowo University, Ile- Ife, Nigeria
8. University of Cape Coast, Cape Coast, Ghana
9. Kwame Nkrumah University of Science and Technology, Kumasi, Ghana
10. University of Abuja, Abuja, Nigeria
11. Parkinson School of Health Sciences and Public Health, Loyola University, Chicago, IL, USA
12. Centre for Research on Genomics and Global Health, National Human Genome Research Institute, National Institutes of Health, Bethesda, MD, USA
13. Division of Nephrology, Boston Medical Center, Boston University School of Medicine, Boston, MA, USA
14. Noguchi Memorial Institute for Medical Research, University of Ghana, Ghana
15. Department of Pediatrics, University of Toronto, Toronto, Canada.
16. Department of Pediatrics, Duke University Medical Center, Durham, NC, USA
17. University of Western Cape, Cape Town, South Africa
18. Harvard Medical School, Harvard University, Boston, MA, USA
19. Nnamdi Azikiwe University Teaching Hospital, Nnewi, Nigeria
20. Aminu Kano Teaching Hospital, Kano, Nigeria
21. Delta State University Teaching Hospital, Warri, Nigeria
22. Lagos State University Teaching Hospital, Lagos, Nigeria
23. Lagos University Teaching Hospital, University of Lagos, College of Medicine, Lagos, Nigeria
24. Usmanu Danfodiyo University Teaching Hospital, Sokoto, Nigeria
25. University of Illinois at Chicago, Chicago IL, USA
26. University of Yaoundé, Yaoundé, Cameroon
27. University of Cape Town, Cape Town, South Africa
28. New York University, New York City, NY, USA
29. Duke University, Durham, NC, USA
30. Technion-Israel Institute of Technology, Haifa, Israel
31. University of Kansas School of Medicine, Kansas City, KS, USA

**Deceased

**Supplementary Figure S1.** Optimal cluster determination and similarity matrices using different clustering algorithms from transcriptomic profiles of MCD and FSGS participants based on kidney biopsy tubulointerstitial gene expression data in NEPTUNE (A), ERCB (B), and H3 Africa (C). Sankey diagrams from the ERCB (D) and H3 Africa (E) cohorts comparing diagnosis with cluster assignment.

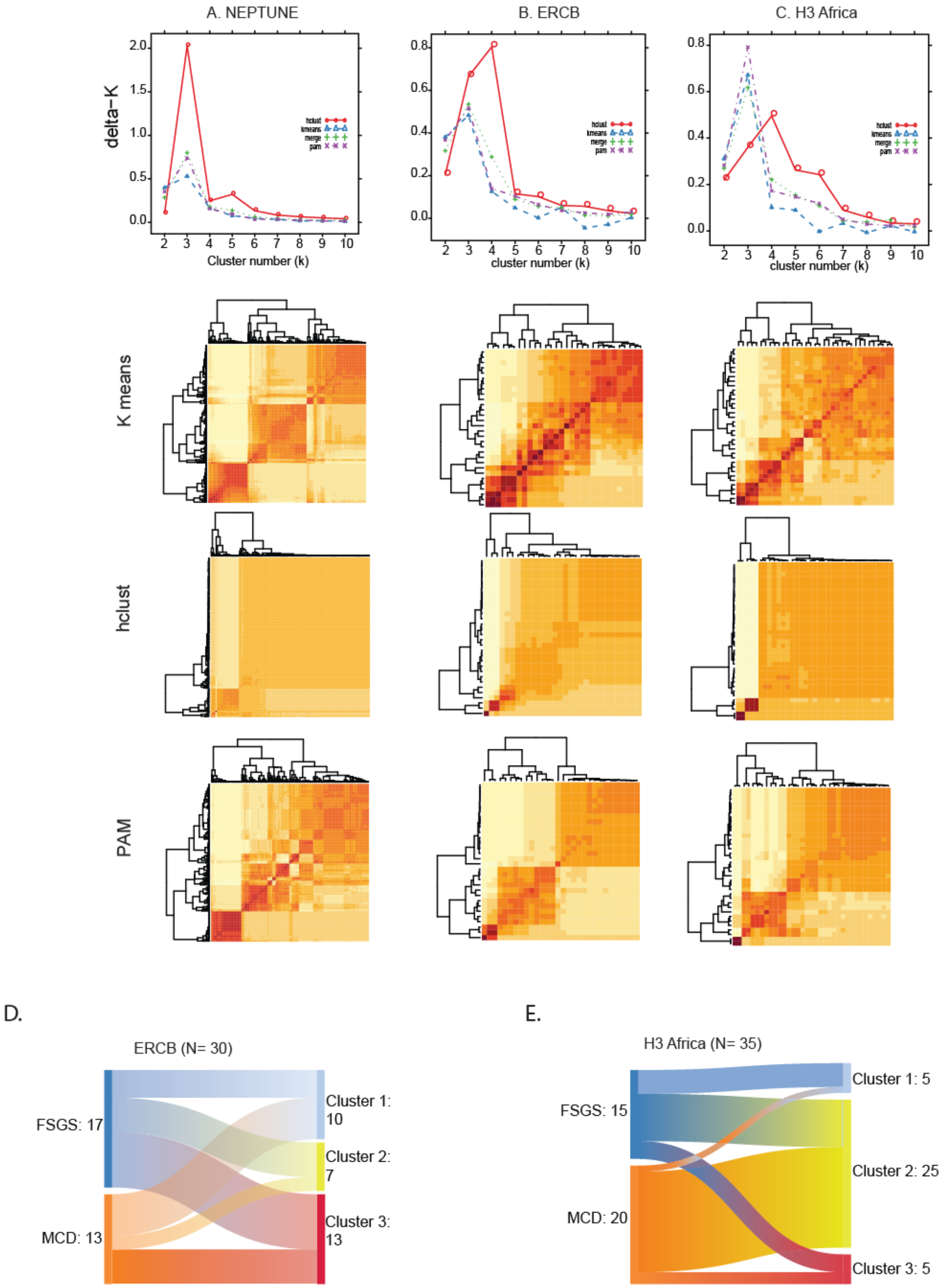

**Supplementary Figure S2** Optimal cluster determination and similarity matrices using different clustering algorithms from transcriptomic profiles of MCD and FSGS participants based on kidney biopsy glomerular gene expression data in NEPTUNE (A), and ERCB (B).

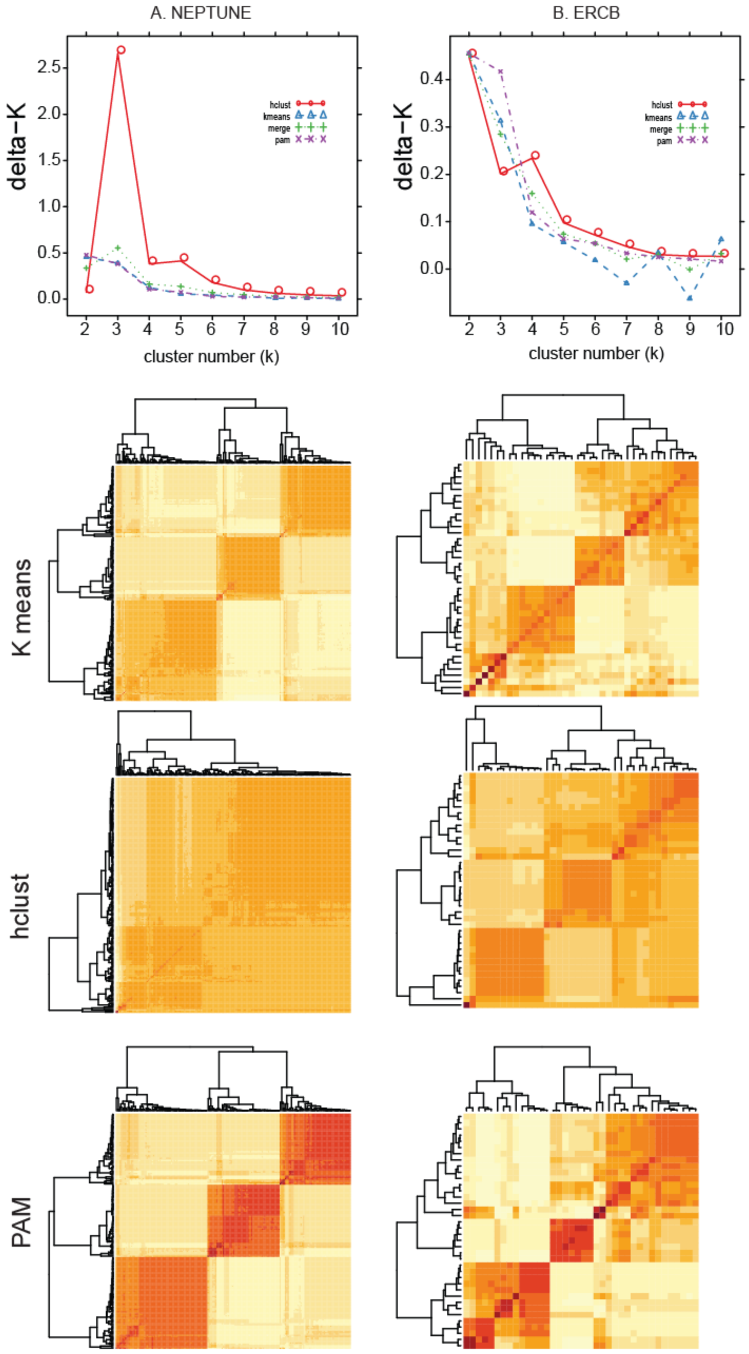

**Supplementary Figure S3.** Clinical information by cluster for eGFR (A), Age (B), and UPCR (C) across the cohorts analyzed.

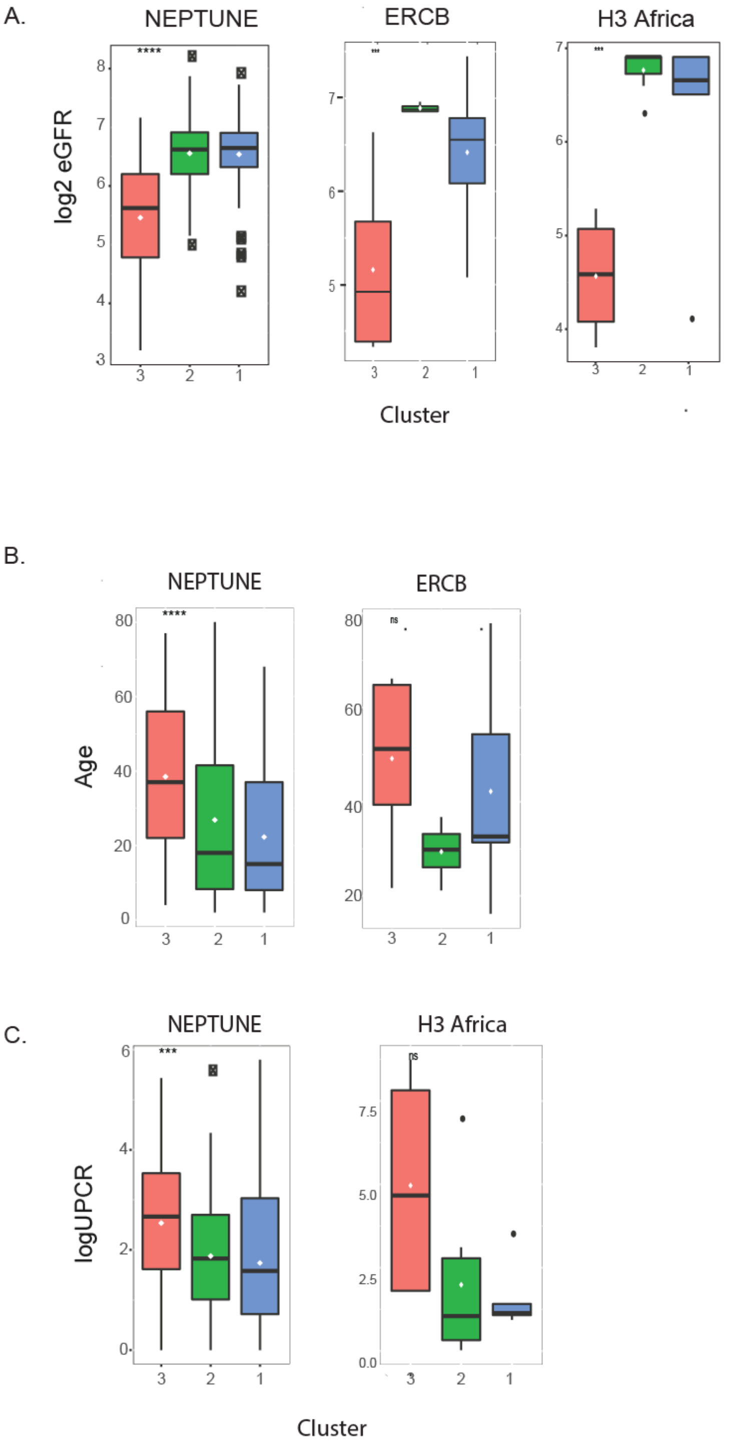

**Supplementary Table S1.** Biological upstream regulators from the differential expression profile in each cohort were extracted from Ingenuity Pathways Analysis and ranked according to enrichment p-value for each upstream regulator in a given cohort. Upstream regulators were aggregated and the average rank and activation score was calculated. The top 15 regulators by average rank are shown.

| Upstream Regulator | Molecule Type | Average Rank | Average Activation Z-score |
|---|---|---|---|
| TNF | cytokine | 1.3 | 11.03 |
| IFNG | cytokine | 2.0 | 9.58 |
| TGFB1 | growth factor | 4.3 | 7.33 |
| IL6 | cytokine | 4.7 | 6.49 |
| IL1B | cytokine | 6.7 | 8.84 |
| STAT3 | transcription regulator | 11.7 | 5.55 |
| SMARCA4 | transcription regulator | 12.7 | 6.75 |
| KRAS | enzyme | 14.7 | -1.50 |
| Immunoglobulin | complex | 15.0 | -0.80 |
| IL4 | cytokine | 15.3 | 3.68 |
| OSM | cytokine | 16.0 | 7.07 |
| NFkB (complex) | complex | 17.7 | 8.42 |
| STAT1 | transcription regulator | 18.0 | 6.71 |
| ERBB2 | kinase | 21.0 | 4.26 |
| TP53 | transcription regulator | 22.0 | 3.92 |

**Supplementary Table S2.** Genes and gene products activated by TNF used to generate the TNF activation score.

| Gen | PMII | Gene | PMID | Gene4 | PMID | Gene | PMID | Gene8 | PMID | Gene1 | PMID1 | Gene12 | PMID1 |
|---|---|---|---|---|---|---|---|---|---|---|---|---|---|
| CCL17 | 12354417 | CHUK | 11796489 | EMCN | 11594763 | ICAM1 | 15963988 | ITGB2 | 12454401 | OLR1 | 11985903 | SYTL1 | 12137562 |
| CCL19 | 10679062 | COX2 | 10501211 | ENG | 17389265 | ICOSLG | 10744980 | JUN | 17500068 | OPTN | 10807909 | TAP1 | 1385520 |
| CCL2 | 9825772 | CPNE1 | 14674885 | EPCAM | 10867614 | IER3 | 11244505 | JUNB | 10903323 | ORM1 | 9726030 | TFRC | 9135559 |
| CCL20 | 11133838 | CR1 | 2961377 | ESAM | 15505101 | IGFBP1 | 10070049 | KIT | 10206577 | OSM | 12097485 | TGFB1 | 15574511 |
| CCL22 | 11923841 | CRH | 12676571 | ETS1 | 11229456 | IGFBP3 | 11971816 | KITLG | 15579452 | PAPPA | 16269458 | THBS1 | 11157717 |
| CCL27 | 11821900 | CRHR2 | 17412781 | F3 | 11058594 | IGFBP6 | 12054123 | LAMP3 | 15963988 | PECAM1 | 7686548 | TIMP1 | 15663564 |
| CCL3 | 12115625 | CRP | 9726030 | FABP2 | 11329616 | IKBKB | 11796489 | LEP | 12032749 | PLA2G4A | 10930295 | TIMP2 | 15663564 |
| CCL4 | 9570566 | CSF1 | 3494061 | FABP4 | 12927809 | IL10 | 12414777 | LIF | 8895217 | PLAT | 12692009 | TLR2 | 11160251 |
| CCL5 | 9277410 | CSF2 | 12562880 | FANCG | 11181053 | IL11 | 10467228 | MADCAM1 | 15483224 | PLAU | 8601416 | TLR4 | 11160251 |
| CCND1 | 12444159 | CSF3 | 1700731 | FAS | 8168998 | IL12B | 10513808 | MAP2K6 | 9029150 | PLAUR | 8601416 | TNF | 16365392 |
| CCR1 | 9787141 | CTSG | 11722574 | FASLG | 10358159 | IL13RA2 | 14652008 | MAP3K14 | 11368442 | PLD1 | 11485559 | TNFAIP3 | 12388275 |
| CCR3 | 11884459 | CX3CL1 | 11525637 | FCER2 | 7643018 | IL15RA | 12165497 | MAP4K4 | 16461467 | POSTN | 15378733 | TNFRSF11B | 11685652 |
| CCR4 | 9787141 | CXADR | 12571626 | FGF2 | 10629075 | IL1A | 7686496 | MEFV | 10807793 | PRTN3 | 11415941 | TNFRSF1A | 11923841 |
| CCR5 | 9787141 | CXCL1 | 12744776 | FGF7 | 7936642 | IL1B | 7994029 | MIPEP | 1372592 | PTGES | 11029586 | TNFRSF1B | 12067756 |
| CCR7 | 15963988 | CXCL10 | 11884459 | FOS | 3259871 | IL1R1 | 8120407 | MMP1 | 12060661 | PTGS2 | 12576525 | TNFRSF21 | 11753679 |
| CCRL2 | 15188357 | CXCL11 | 12627325 | GADD45B | 12388275 | IL1RN | 8331299 | MMP10 | 8349617 | PTHLH | 10854575 | TNFRSF8 | 12414777 |
| CD14 | 1373513 | CXCL2 | 10502561 | GDF15 | 9886240 | IL2 | 12414777 | MMP12 | 17525194 | PTPRF | 10905491 | TNFSF10 | 12218071 |
| CD1A | 11238627 | CXCL3 | 9277410 | GFAP | 10674496 | IL21R | 11986233 | MMP13 | 12878172 | PTX3 | 9521058 | TNFSF11 | 15479886 |
| CD4 | 1921451 | CXCL5 | 9277410 | GJA1 | 12827213 | IL2RA | 12414777 | MMP14 | 9212749 | RB1 | 15611081 | TNFSF15 | 11911831 |
| CD40 | 11488834 | CXCL6 | 12744776 | GNAI3 | 12214898 | IL3RA | 1825289 | MMP17 | 12962706 | RELA | 9756499 | TNIP2 | 15378733 |
| CD55 | 10477692 | CXCL8 | 9209275 | GPNMB | 19320736 | IL6 | 10467228 | MMP2 | 25358651 | RPS6KA5 | 9873047 | TP53 | 10213465 |
| CD58 | 1354203 | CXCL9 | 12627325 | GRIA1 | 12460558 | IL6R | 11923841 | MMP3 | 10967046 | S100A11 | 16339570 | TRAF1 | 11978013 |
| CD70 | 9067541 | CXCR4 | 11160334 | GSS | 15378733 | IL7 | 10961889 | MMP7 | 9461124 | SAA1 | 1714101 | TRAF2 | 12067756 |
| CD80 | 9824486 | CYCS | 12149248 | HBEGF | 12138120 | IL7R | 10779425 | MMP9 | 25358651 | SELE | 8695814 | TRAF3 | 12067756 |
| CD83 | 15963988 | CYLD | 18245814 | HCK | 12237848 | INHBA | 1417851 | MUC1 | 10867614 | SELL | 12517920 | TRAF4 | 12067756 |
| CD86 | 12230943 | CYP19A1 | 12452446 | HGF | 9397161 | IRF1 | 12713595 | MUC5AC | 12690113 | SELP | 10484438 | TRAF5 | 12067756 |
| CDK3 | 15378733 | DDR1 | 11606478 | HIF1A | 12808024 | ITGA1 | 12606473 | MYC | 10200535 | SERPINA3 | 9461605 | TRAF6 | 12067756 |
| CDK4 | 15611081 | DEFB4A | 11702237 | HLA-A | 1354203 | ITGA4 | 12759453 | MYLK | 15681825 | SERPINB2 | 8824249 | TRPC1 | 12855710 |
| CDKN1A | 12795334 | DUSP10 | 24707477 | HLA-B | 3455781 | ITGA5 | 8006453 | NAMPT | 11241162 | SERPINE1 | 11402043 | TXN | 10555039 |
| CEBPA | 10211885 | E2F1 | 15611081 | HLA-C | 1354203 | ITGA6 | 8006453 | NFKB1 | 9756499 | SLC11A2 | 16224057 | TYMP | 14573775 |
| CFB | 7512988 | EDN1 | 10764953 | HMOX1 | 10330231 | ITGAL | 10198263 | NFKB2 | 9529131 | SLC3A2 | 2318252 | VCAM1 | 10484438 |
| CFH | 1690734 | EFNA1 | 11278471 | HP | 9097927 | ITGAM | 12747235 | NFKBIA | 12580918 | SLC7A2 | 11742806 | WNT5A | 12165812 |
| CFLAR | 12861043 | EGFR | 10077640 | HPSE | 16384929 | ITGAX | 1921451 | NLRP3 | 14662828 | SMAD7 | 10652273 | XIAP | 16219905 |
| CHRM2 | 15671275 | EGR1 | 1370482 | HSP90B1 | 15378733 | ITGB1 | 9565575 | NOS2 | 10601128 | SOD2 | 9114748 | ZFP36 | 10763822 |

**Supplementary Table S3.** Clinical characteristics of patients with kidney biopsies profiled by snRNAseq

| | TNF High (N = 5) | TNF Low (N = 5) |
|---|---|---|
| Age mean (sd) | 44.6 (22.6) | 20.8 (17.4) |
| Sex (male:female) | 2:3 | 3:2 |
| eGFR mean (sd) | 44.26 (22.47) | 115.53 (44.01) |
| UPCR median (IQR) | 5.05 (3.62, 5.8) | 0.96 (0.86, 1.03) |
| FSGS | 4 | 4 |
| Cluster 3 | 5 | 0 |
| TNF Score mean (sd) | 0.13 (0.02) | 0.081 (0.01) |
